# LZTR1 functions as a two-hit tumor suppressor in childhood acute lymphoblastic leukemia

**DOI:** 10.64898/2026.06.26.26356641

**Authors:** Adeline A. Bonnard, Aurélie Caye-Eude, Chloé Arfeuille, Séverine Drunat, Anna Dehler, Fabio D. Steffen, Elodie Lainey, Damien Bodet, Claire Freycon, Catherine Paillard, Pauline Simon, Arnaud Petit, Cécile Pochon, Jean-Hugues Dalle, Nastassja Scheidegger, Beat Bornhauser, André Baruchel, Marion Strullu, Yoann Vial, Hélène Cavé

## Abstract

LZTR1 negatively regulates RAS family proteins via proteasomal degradation. Germline loss-of-function variants cause Noonan syndrome, with emerging evidence implicating LZTR1 in predisposition to childhood acute lymphoblastic leukemia (ALL), though its role in hematopoiesis remains poorly defined. Screening 1,587 children with ALL identified *LZTR1* variants in 44 patients (2.8%). Germline variants were detected in 32 patients (2.0%), a frequency comparable to that observed in the general population (1.75%; 1,925/110,017; p=0.50). Somatic *LZTR1* alterations were identified in 22 patients (1.4%) and were predominantly bi-allelic, arising through either a germline-plus-somatic or dual somatic configuration. They persisted at relapse. Despite enrichment in favorable-risk subtypes (*ETV6::RUNX1*, high-hyperdiploid, ERG/DUX4), bi-allelic *LZTR1*-mutated cases showed delayed minimal residual disease clearance and higher late relapse risk, identifying a subgroup unsuitable for treatment de-escalation. LZTR1 expression was increased in most wild-type leukemias, consistent with a compensatory response to aberrant RAS pathway activation. Bi-allelic *LZTR1* inactivation abolished RAS regulation, leading to deregulated canonical RAS expression and ectopic expression of the non-canonical RIT1 protein, whose involvement in ALL has not previously been reported. These findings establish LZTR1 as a classical tumor suppressor in ALL via a two-hit model. Monoallelic alterations show insufficient signaling perturbation and low germline penetrance, whereas bi-allelic inactivation acts as a driver event linked to a high risk of late relapse despite favorable genomics.

## Introduction

Acute lymphoblastic leukemia (ALL) is the most common malignant tumor in children. It encompasses a variety of different entities, each characterized by distinct somatically acquired recurrent driver genetic alterations assumed to initiate the leukemia, shape its biology and influence outcome(1). These lesions are fundamental to the classifiers on which risk-based therapy relies. For instance, in B-cell progenitor ALLs (BCP-ALL), the presence of high hyperdiploidy (HeH), *ETV6::RUNX1* fusion, or *ERG* deletion associated with a *DUX4* rearrangement (*ERG/DUX*) defines good-risk leukemia that can be successfully treated with low-intensity therapy(2–8).

Alongside these ‘classifying’ genetic lesions, a number of conditions have been shown to influence leukemia susceptibility across a wide range of penetrance levels(9). One such condition is Noonan syndrome (NS), a developmental disorder caused by germline genomic variations, which activate the RAS/MAPK signaling pathway. NS has been shown to preferentially increase the risk of childhood BCP-ALL of the high hyperdiploid subtype(10).

LZTR1 (Leucine zipper-like transcriptional regulator 1), an adaptor protein involved in CUL-3-based ubiquitin ligase complex, has recently emerged as a key post-translational regulator of the RAS pathway. It acts by promoting the ubiquitination and subsequent degradation of RAS family GTPases by the proteasome, thereby preventing excessive activation of oncogenic signalling(11–13). LZTR1 contains six Kelch-repeat motifs in its N-terminal region, which mediate substrate recognition, and two BTB (Back-to-Back) domains in its C-terminal region, which facilitate homodimerization and interaction with CUL3 for ubiquitination complex assembly. *LZTR1* is increasingly recognized as a tumor suppressor gene with somatic mutations identified in glioblastoma(14).

Germline inactivating mutations in *LZTR1* (22q11.2) were initially associated with schwannomatosis(15), and, more recently, with NS of either autosomal dominant (AD) or recessive (AR) inheritance(16,17). In AD-NS, mutations are restricted to the Kelch domains and exert a dominant-negative (DN) effect. In contrast, AR-NS mutations are distributed across the entire protein and typically involve one hypomorphic allele paired with a null allele(17).

Intriguingly, among the few documented cases of NS with *LZTR1* mutations (LZTR1-NS), four individuals developed childhood ALL(17–19), suggesting that patients carrying germline *LZTR1* mutations may be at risk of developing ALL. Germline variants in *LZTR1* are among the most frequently reported in the St. Jude Children’s hospital database and a predisposition to developing HeH BCP-ALL has been proposed in patients with germline mutations(20).

This study investigated the genetic alterations of *LZTR1* in childhood ALL and the consequences of *LZTR1* inactivation in this disease, including which RAS proteins are targeted.

## Patients and Methods

### Patients

The study cohort included 1587 children with newly-diagnosed ALL (1380 BCP-ALL; 207 T-ALL) consecutively referred to our laboratory between 2018 and 2023. Patients were treated according to the EORTC-58081 (NCT01185886), FRALLE, CAALLF_01 (NCT02716233) or EsPhALL (NCT00287105) trials. Written informed consent was obtained from patients or parents according to national regulations. The study was approved by the institutional review board of “Hôpitaux Universitaires Paris-Nord Val-de-Seine” (IRB-00006477) and conducted in accordance with the Declaration of Helsinki.

### Population frequency study

General population data were obtained from the non-cancer subset of GnomAD v2.1.1 (January 13, 2023)(21). Variant filtering followed Deng *et al* (2022)(22) using the canonical *LZTR1* transcript (ENST00000215739.8), with minor adjustments (*Supplementary Information*).

### Genetic screening

Genomic DNA and RNA were extracted from diagnostic, remission, and/or relapse samples. Karyotype and multiplex ligation-dependent probe amplification (MLPA) (MRC-Holland SALSA® MLPA® Probemix P335) were part of the diagnostic workup. Mutations were screened using whole-exome sequencing (WES) and/or targeted next-generation sequencing (NGS) (gene list and bioinformatic details in *Supplementary Information*). Variants with a variant allele frequency (VAF) <2 % or a population frequency ≥5×10⁻L in GnomAD v2.1.1 (non-cancer) were excluded. Germline status was determined using matched remission samples. Complete sequencing of the *LZTR1* locus (including 5’/3’UTR) was performed on samples displaying RNA imbalance. Somatic variants were classified according to AMP/ASCO/CAP guidelines(23). Germline variants were classified according to ACMG/AMP guidelines(24).

Genome-wide copy-number was evaluated by Illumina OmniExpress 700K SNP-array. Copy neutral loss of heterozygosity (CN-LOH) was identified using GenomeStudio (Illumina) and/or NGS-based genotyping.

### RNA sequencing

Whole-transcriptome RNA sequencing (RNAseq) was performed using the NEBNext Ultra II Directional RNA Library Prep Kit (New England Biolabs) and NovaSeq paired-end technology (Illumina) on the IntegraGen platform (Evry, France). Targeted RNAseq was performed using the SureSelect XTHS2 RNA kit (Agilent Technologies) on a NextSeq 500 system (Illumina).

Expression quantification and splice junction analyses were performed using standard pipelines. Unsupervised and differential expression analyses were conducted with the Galileo platform (IntegraGen).

### Cell culture and CRISPR/Cas9 knock-out

The cell lines used are described in Table S1. Gene inactivation was achieved using Cas9-GFP plasmids (Sigma-Aldrich) carrying gRNAs for *LZTR1* or the AAVS1 control locus (*PPP1R12C*). Transfections were performed with a Lonza-4D-Nucleofector. Single-cell clones were isolated by flow cytometry and validated by Sanger sequencing and capillary immunoassay after 14-day culture. WES was performed to check for off-target effects.

Signaling studies were performed after 24-hour serum deprivation (1% FBS). Cells were then stimulated by recombinant GM-CSF (TF-1) or 10% FBS (Jurkat). Detailed cloning, nucleofection, culture and stimulation protocols are in *Supplementary Information*.

### Protein studies

Protein identification and quantification were conducted by capillary immunoassay using a Jess® automated capillary electrophoresis system (ProteinSimple).

RAS-GTP pull-down assays were performed from total protein extracts using the GST-Raf1-RBD pull-down assay (Thermo Fisher Scientific, #16117).

### Patient-Derived Xenografts (PDX)

To generate PDXs, primary ALL cells were transplanted into NOD/scidIL2Rγnull (NSG) mice mice via tail vein as described previously(25). Engraftment was monitored by flow cytometry, and ALL cells were collected from spleen and bone marrow. All animal experiments were conducted in accordance with institutional and national guidelines for animal care and use, and were approved by the Zurich veterinary office (protocol ZH168/21).

### Drug Response Profiling (DRP)

DRP of spleen-derived PDX cells was performed as reported(26,27). Mesenchymal stromal cells (hTERT-immortalized) were plated for 24 h before adding 10,000 PDX cells. Drugs were added in five-point serial dilutions (Table S2). After 72 h of incubation, viability was measured by CyQUANT staining and automated fluorescence microscopy. Cell numbers were normalized to DMSO, fitted to a three-parameter dose-response model(28), and integrated to obtain logarithmic area-under-the-curve (logAUC) values as a metric for drug sensitivity.

### Statistical analysis

Statistical analyses were performed using GraphPad Prism v10.0 and R. Two-tailed t-tests or ANOVA were applied as appropriate, with p < 0.05 considered significant. Detailed statistical methods are provided in *Supplementary Information*.

## Results

### Germline *LZTR1* variants in children with ALL

Sequencing of leukemia DNA from 1,587 patients (1380 with B-cell precursor ALL, BCP-ALL; 207 with T-cell ALL, T-ALL), identified 51 *LZTR1* variants in 44 patients (2.8%), including 41 BCP-ALL and 3 T-ALL cases (Table 1; Table S3; Figure S1).

**Table 1:**
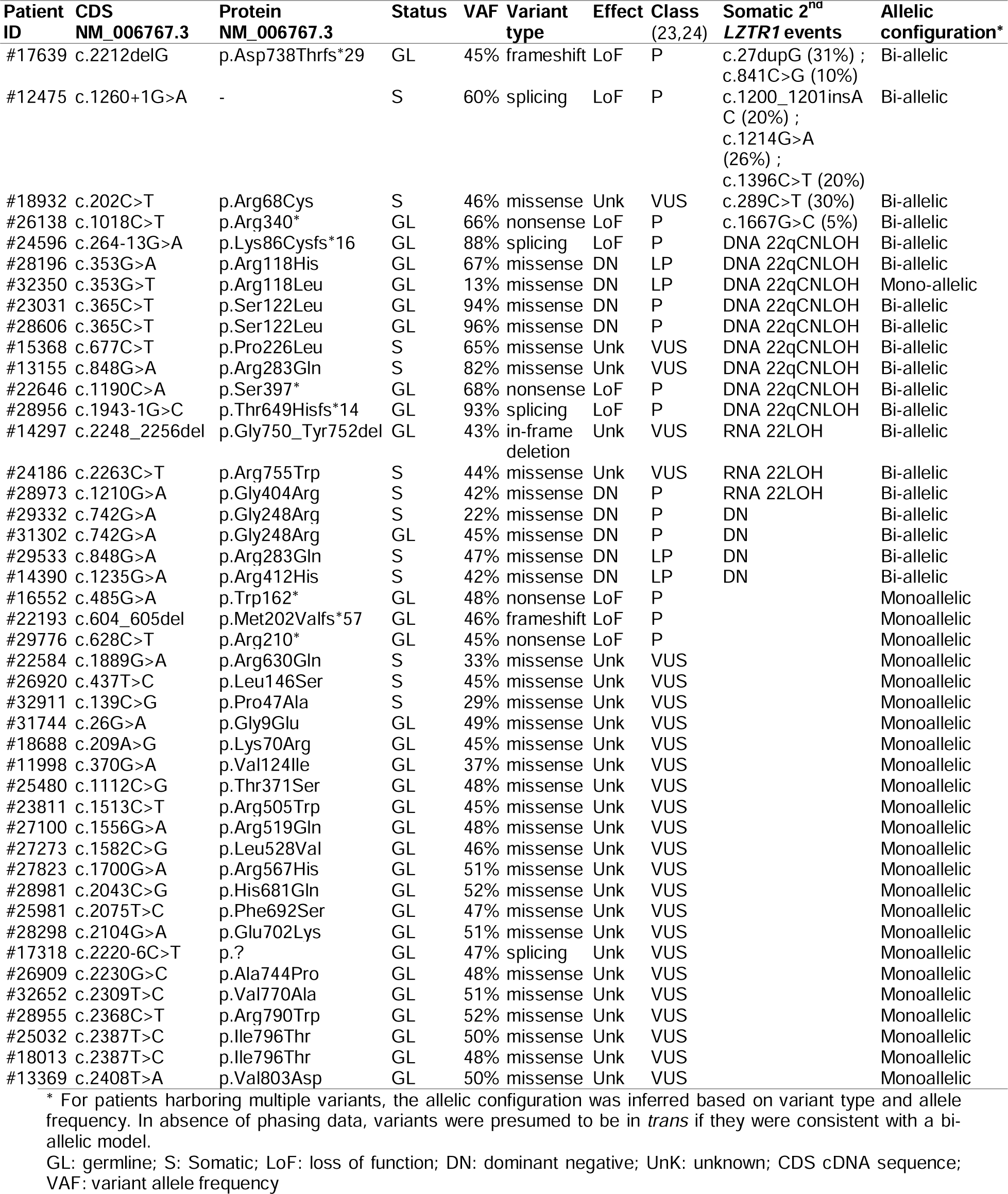
Summary of identified *LZTR1* variants and their pathogenic classification at variant and patient levels * For patients harboring multiple variants, the allelic configuration was inferred based on variant type and allele frequency. In absence of phasing data, variants were presumed to be in *trans* if they were consistent with a bi-allelic model. GL: germline; S: Somatic; LoF: loss of function; DN: dominant negative; UnK: unknown; CDS cDNA sequence; VAF: variant allele frequency.

Thirty-two of the 51 variants, were confirmed germline based on their presence in non-tumoral samples (Table S3). GnomAD v2.1.1 non-cancer controls showed *LZTR1* variant prevalence of 1.75% (1,925/110,017), with missense tolerance (o/e = 0.93; 95% CI 0.86–1.0) and truncating enrichment (o/e = 2.28; 95% CI 1.75–1.99; Figure S2A). No bi-allelic deleterious variants co-occurred in GnomAD, indicating intolerance to complete LZTR1 loss.

Germline *LZTR1* variant frequency in ALL patients (32/1,587; 2.0%) was not significantly elevated versus population rates (1.75%; p = 0.50), mirroring background prevalence.

These findings demonstrate that while both germline and somatic *LZTR1* variants occur in ALL, germline variants are not enriched compared to the general population.

### *LZTR1* is a leukemogenic driver in children with ALL

Among 51 *LZTR1* variants identified, 11 truncating variants (including 2 splice; Figure S3) and 5 germline missense/indel variants were classified pathogenic/likely pathogenic (P/LP) (Figure 1A; Table S3). The dominant-negative p.Gly248Arg variant, recurrent in autosomal dominant Noonan syndrome (AD-NS)(11), was identified in the single patient with clinical NS. Other germline missense variants, predominantly C-terminal (14/17), were variants of uncertain significance (VUS)(29) (Figure 1; Table 1, Table S3). Somatic missense variants (n=16) were classified P/LP (Figure 1B, Table 1). P/LP variant distribution mirrored autosomal recessive NS and schwannomatosis patterns, except for the DN variant (Figure S2B-D).

**Figure 1:**
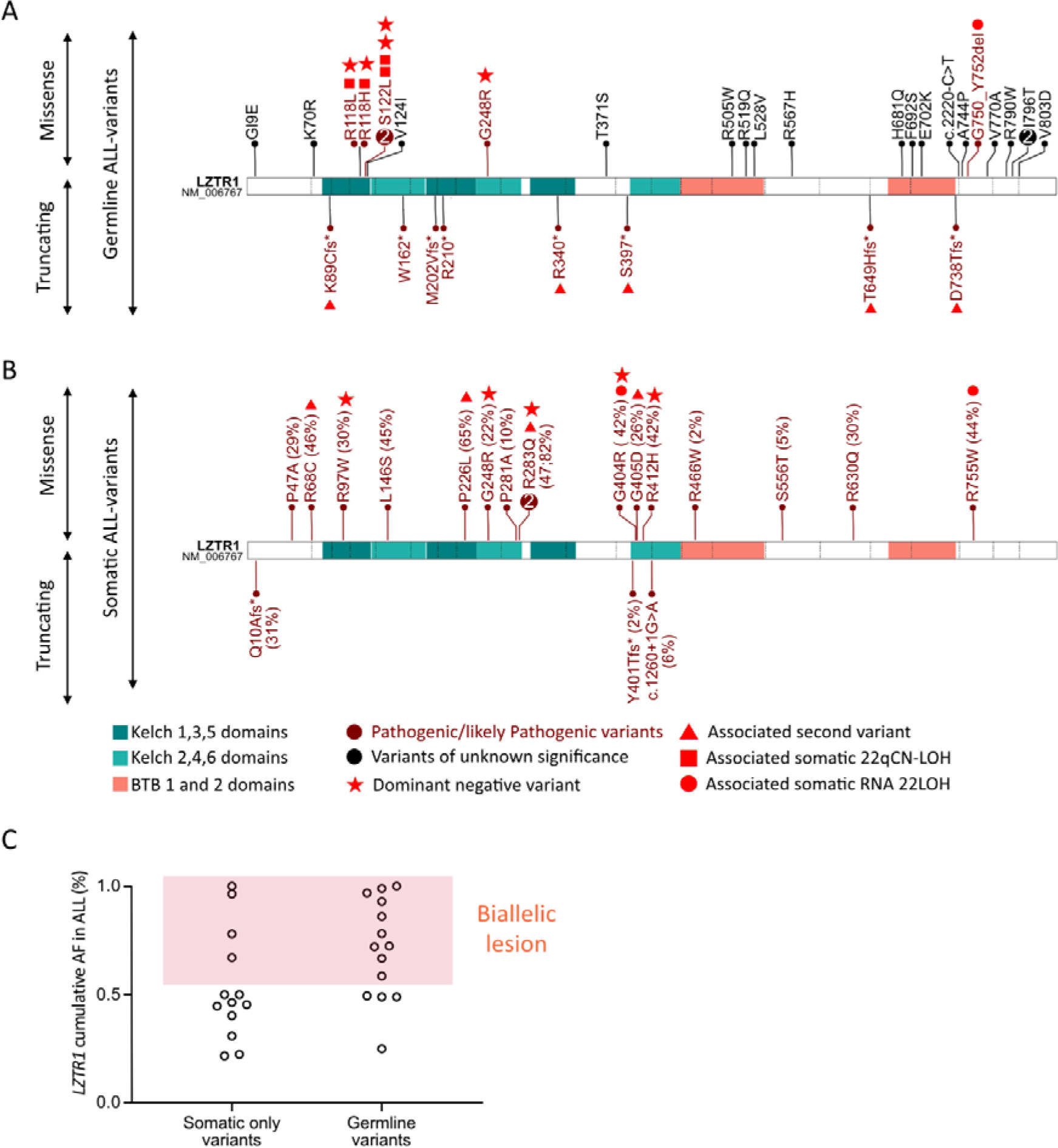
Distribution and allele frequencies of 51 *LZTR1* variants in ALL patients. **(A–B)** Germline **(A)** and somatic **(B)** *LZTR1* variants mapped onto the protein structure, highlighting Kelch (1–6) and BTB (1–2) domains. In both panels, missense variants are shown above and truncating variants below the protein schematic. Numbers inside the lollipops indicate the number of unrelated ALL cases carrying the same variant. For somatic variants (B), the variant allele frequency (VAF, %) is indicated in parentheses next to each variant. Red symbols denote variants associated with at least one additional somatic alteration involving *LZTR1* (e.g., copy neutral loss of heterozygosity (CN-LOH) or RNA-level loss of heterozygosity (RNA LOH)). **(C)** Leukemic cell VAF in patients with somatic-only or germline variants. VAF were adjusted for blast percentage. Dots represent individual patients; multiple variants were summed per patient. A cumulative VAF >0.5 indicates bi-allelic *LZTR1* alteration.

Allelic imbalance was observed in 10 of 44 (23%) *LZTR1*-mutated ALL (*LZTR1*^mut^-ALL) (8/32 germline, 2/12 somatic-only) (Figure 1C; Table 1). SNP-array analysis revealed chromosome 22q copy-neutral loss of heterozygosity (22qCN-LOH) in 9/10 cases (Table S4; Figure S4). 22qCN-LOH has not previously been reported in ALL and was absent among 100 *LZTR1* wild-type controls (*LZTR1*^wt^). One patient with HeH ALL, presented trisomy 22 that duplicated the germline-*LZTR1* allele, plus somatic mutation. Chromosome 22 loss in low-hypodiploid ALL (n=8) occurred exclusively in the carrier of a germline-*LZTR1* variant (TableLS5).

Whole exome sequencing excluded mutations in *VPREB1*, *CHEK2* or other potential targets within 22q11.2 (Table S4). *VPREB1* deletions identified in 4/7 *LZTR1*^mut^ versus 11/29 *LZTR1*^wt^ ALL, postdatedLCN-LOH (Figure S5). Together these findings implicate *LZTR1* as the CN-LOH target.

RNA-seq analyses for transcriptional or splicing anomalies identified LOH at the RNA level (RNA LOH), defined as >25% RNA/DNA VAF imbalance, in 3/34 evaluable cases (1 germline, 2 somatic; Figure S6A-B). RNA-LOH resolved at remission, consistent with somatic silencing of the *LZTR1^wt^* allele. These 3 cases were considered as harboring bi-allelic *LZTR1* inactivation. No underlying DNA alteration was detected by comprehensive genomic sequencing. No RNA-LOH was found in 149 *LZTR1^wt^*-ALL controls (Figure S6B).

Overall, *LZTR1* P/LP bi-allelic inactivation, via secondary variants, DNA or RNA 22qCN-LOH, or DN variants, was documented in 19/1587 patients (1.2%) of whom 10 (53%) carried an initial germline alteration and 9 (47%) had exclusively somatic alterations (Figure 1, Figure S1, Table 1).

Nine of these 19 patients were also studied at relapse (Figure 2). Secondary *LZTR1* alterations persisted in 7/9 cases at first and second (n=2) relapse. In one case, the 22qCN-LOH clone identified at diagnosis was replaced at relapse by a new clone harboring a truncating *LZTR1* variant (#22646), supporting the dependence of leukemia cells on LZTR1 inactivation. The final case showed *LZTR1*-mutated subclones outcompeted by pre-existing RAS-pathway clones (#12475).

**Figure 2:**
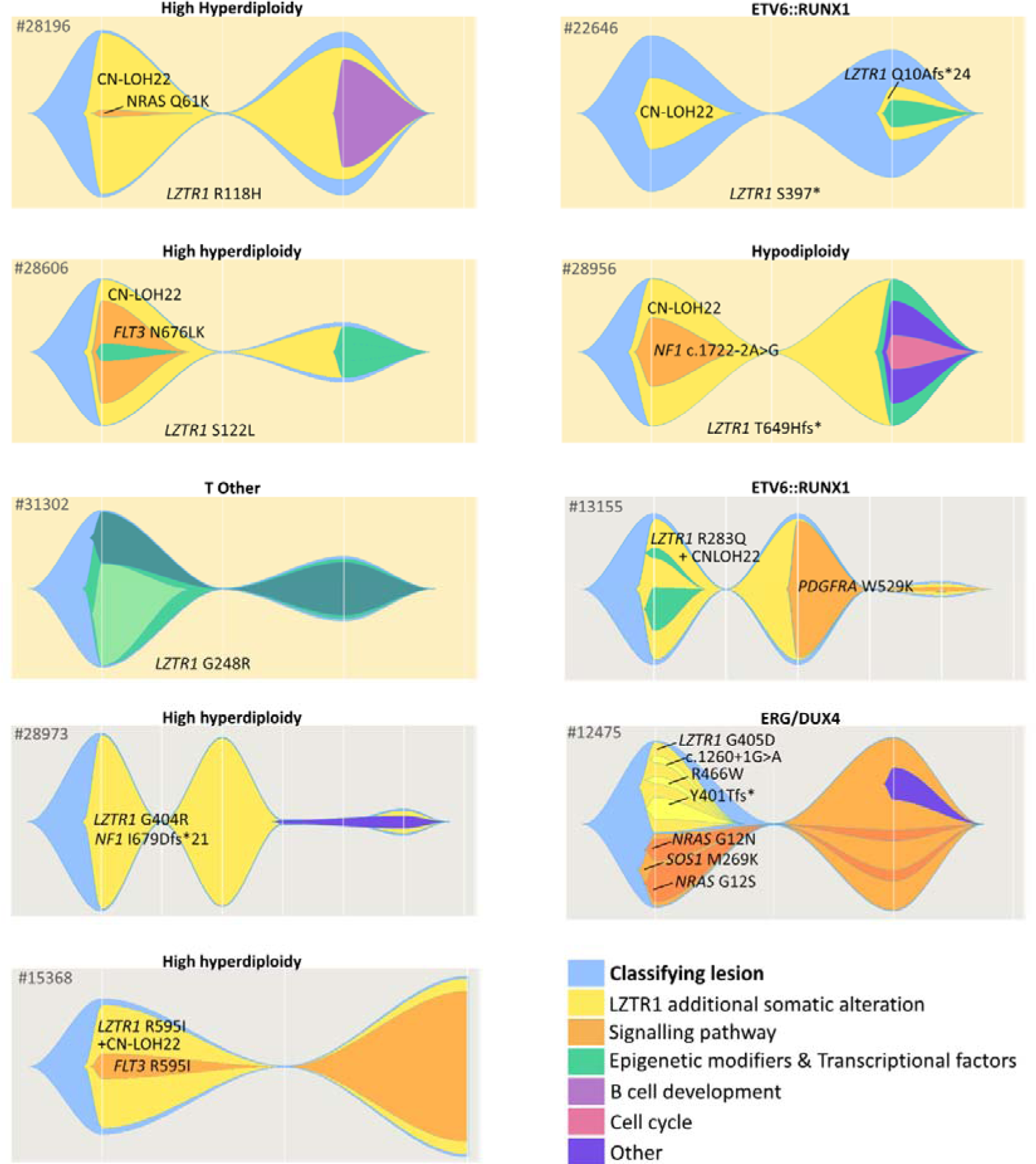
Clonal architecture and evolution in relapsed ALL with *LZTR1* alterations. Fishplots depict clonal composition and dynamics across disease progression in nine relapsed *LZTR1*^mut/mut^-ALL patients. The classifying lesion is shown in blue and labeled above each fishplot. Orange represents *RAS* signaling–related clones, while all other subclones are colored by functional category, as defined in the key. Background color encodes patient *LZTR1* status: yellow: germline variant carriers (exact variant labeled below the fishplot), gray: patients with a somatic *LZTR1* variant.

The non-random association of somatic *LZTR1* alterations with persistence at relapse establishes LZTR1 as a leukemogenic driver, independent of initial germline or somatic origin.

### *LZTR1* alterations are associated with various ALL-classifying lesions

We compared patients harboring *LZTR1* bi-allelic P/LP alteration (*LZTR1*^mut/mut^-ALL; n=19) with those carrying wild-type *LZTR1* (*LZTR1*^wt^-ALL; n=1543) (Table1; Figure 3).

**Figure 3:**
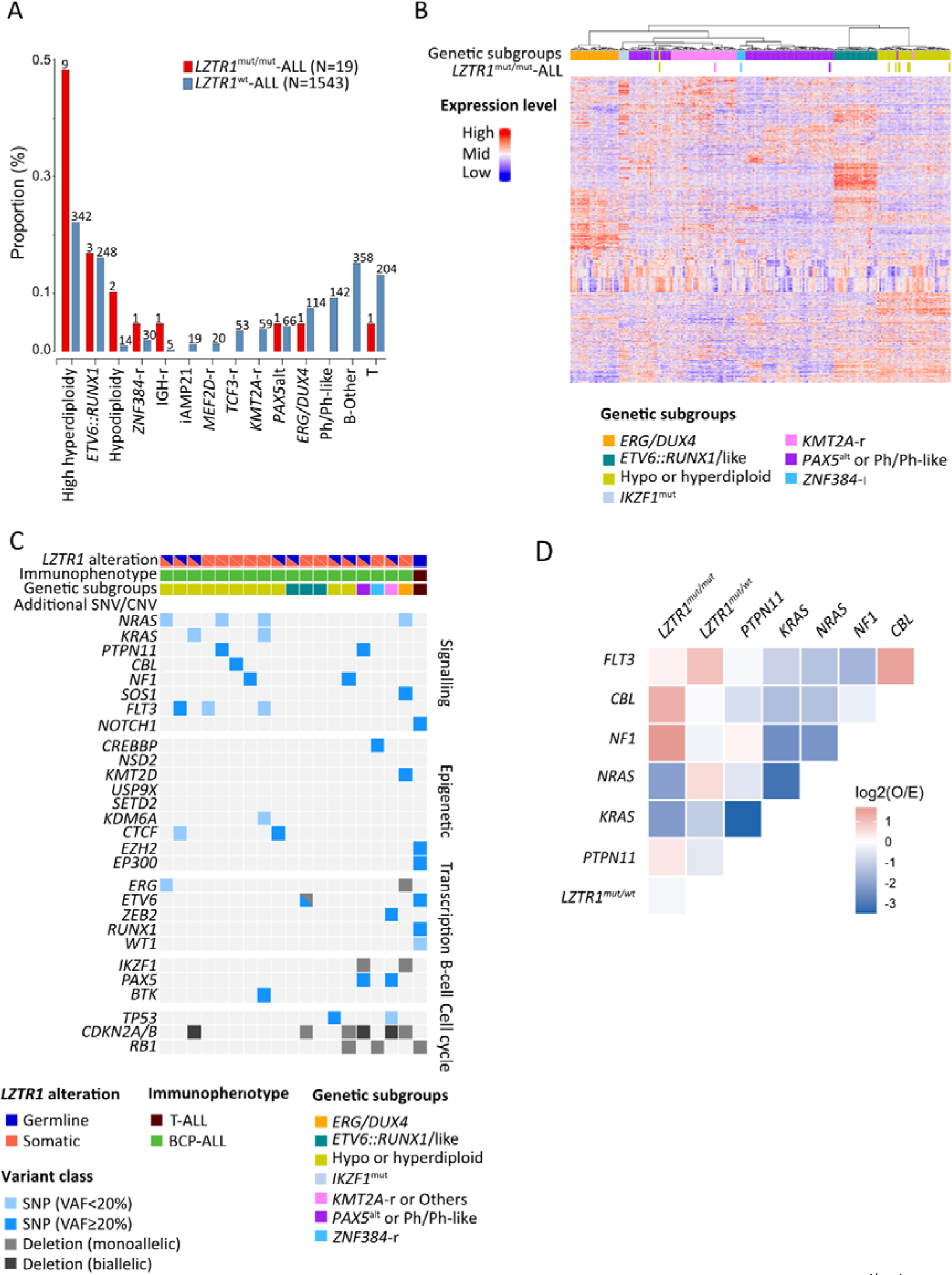
Patients and ALL features of bi-allelic *LZTR1*-mutated ALL. (*LZTR1*^mut/mut^-ALL) **A.** Classifying lesion in *LZTR1* mutated (*LZTR1*^mut/mut^-ALL) versus *LZTR1* wild-type (*LZTR1*^wt^-ALL) **B.** Heatmap showing the unsupervised hierarchical clustering of *LZTR1*^mut/mut^-ALL samples (n=10) and LZTR1^wt^-ALL (n=204) BCP-ALL according to transcriptomic data. **C.** Classifying genetic alterations and secondary single nucleotide alterations (SNA) and copy-number alterations (CNA) identified in ALL with *LZTR1* variants. **D.** Heatmap of pairwise co-mutation enrichment among RAS pathway genes (*KRAS, NRAS, PTPN11, NF1, FLT3, CBL,* and *LZTR1*) in the cohort comparing patients with bi-allelic *LZTR1* pathogenic/likely pathogenic alterations (*LZTR1*^mut/mut^-ALL; n = 19) and patients with wild-type *LZTR1* (*LZTR1*^wt^-ALL; n = 1543). Mutations were defined based on cumulative VAF > 20%. Expected co-mutation frequencies were estimated under an independence model. Negative values (blue) indicate mutual exclusivity, whereas positive values (red) indicate co-occurrence.

Median age at diagnosis was comparable between *LZTR1*^mut/mut^-ALL and *LZTR1*^wt^-ALL (Table 1). Patients with germline alterations were older than those with somatic alterations (mean 8.2 vs 6.9 years, paired t-test, p = 0.02). Genetic subtype distribution differed significantly (p=0.037, Table 1), with HeH BCP-ALL enriched in *LZTR1*^mut/mut^-ALL (9/19; 47%) versus *LZTR1*^wt^-ALL (342/1,367; 25%) (Figure 3A). Other BCP-ALL subtypes included *ETV6::RUNX1* (n=2), hypodiploid (n=2), *ERG/DUX4*, Ph/Ph-like, *PAX5*^alt^, *ZNF384*-r, and *IGH*-r (one case each).

Transcriptome analysis showed *LZTR1*^mut/mut^-ALL clustering by classifying alteration, without a distinct *LZTR1*-specific signature within subtypes (Figure 3B, Figure S6C). Similarly, the pattern of secondary alterations reflected the classifying genotype rather than *LZTR1* status (Figure 3C; Figure S7). Concurrent RAS pathway mutations occurred in 13/26 *LZTR1*^mut/mut^-ALL (*N/KRAS* n=8, *PTPN11* n=3, *CBL* n=2, *NF1* n=2, *SOS1* n=1), often subclonal (n=5) (Figure 2, Figure S7). Unlike alterations affecting RAS pathways regulators, *NRAS* and *KRAS* variants detected in *LZTR1*^mut/mut^-ALL were never carried by the dominant clone, indicating mutual exclusion at the clonal level (Figure 2-3D).

### *LZTR1* alterations identifies poor outcome cases within good-risk ALL patients

Despite enrichment of good-risk BCP-ALL subtypes (*ETV6::RUNX1*, HeH, or *ERG/DUX4* in 63% versus 46% of the *LZTR1^wt^*-ALL; p=0.10) (Table 2), *LZTR1*^mut/mut^-ALL showed slower MRD clearance (Table 3). Relapse occurred more frequently (47.4% vs 15.9%; χ² p=0.001) and later in *LZTR1*^mut/mut^-BCP-ALL, with a median time to relapse of 4.0 years vs 2.4 years in *LZTR1^wt^*-ALL (Table 2-3; Figure 4B). The 5-year cumulative incidence of relapse (CIR) reached 33.2%, in *LZTR1*^mut/mut^-BCP-ALL compared with 16.8% in *LZTR1^wt^*-ALL. Accordingly, EFS dropped steeply after 5 years in *LZTR1*^mut/mut^-BCP-ALL (Figure 4B).

**Figure 4:**
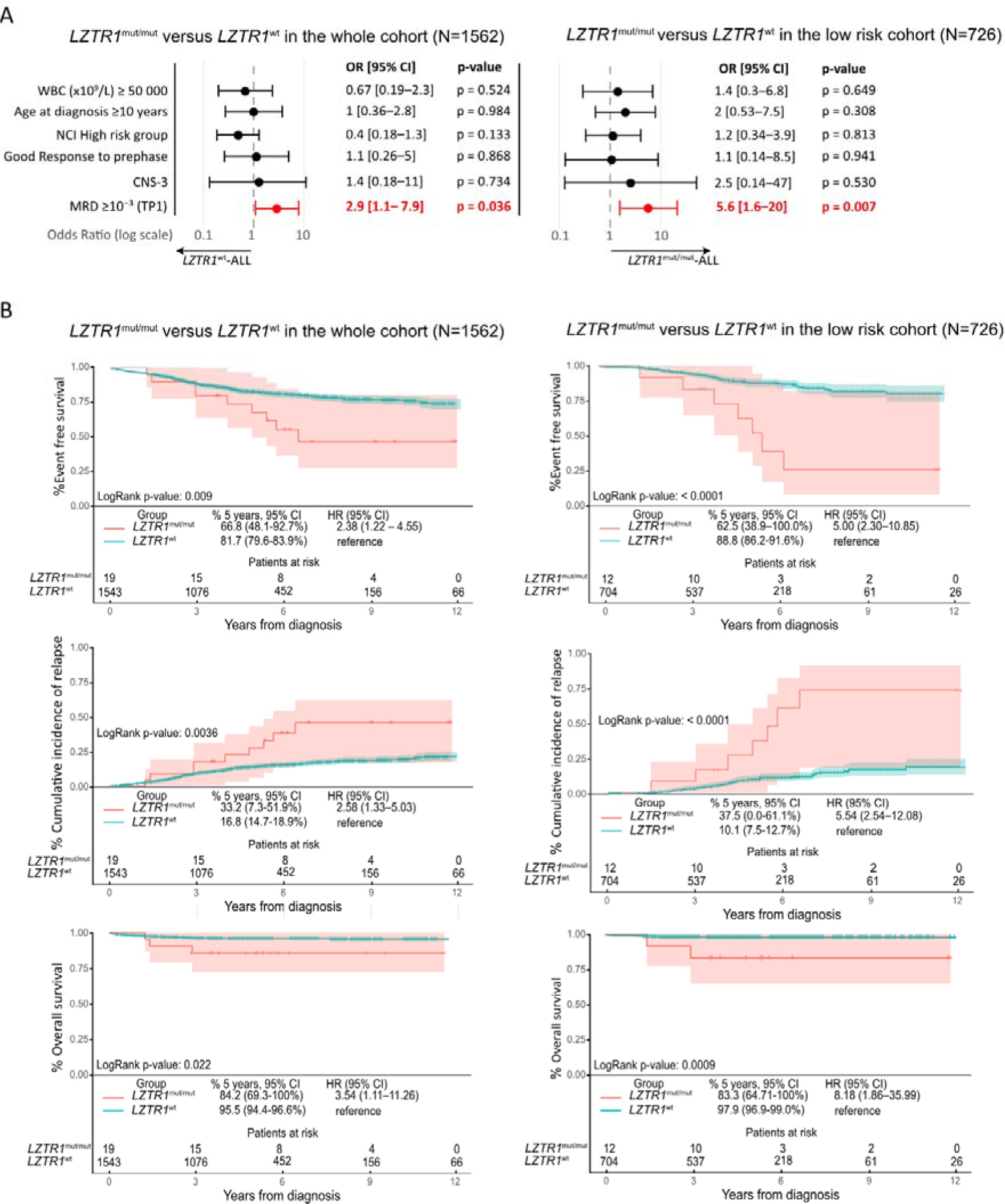
Outcome of patients with bi-allelic *LZTR1*-mutated BCP-ALL. **A.** Graphical representation of odds rations (OR) with 95% confidence intervals (CI). **B.** Event-free survival, cumulative incidence of relapse and overall survival in patients with BCP-ALL harboring *LZTR1* mutations (*LZTR1*^mut/mut^-ALL) versus wild-type (*LZTR1*^wt^-ALL) in the overall cohort (left panels) and the genetic good-risk only subgroup (*ETV6::RUNX1*, High Hyperdiploidy, *ERG/DUX4*) (right panels).

**Table 2:** Patient characteristics according to *LZTR1* status in the overall cohort. Patients with VUS (n=18) or monoallelic *LZTR1* variant (n=6) were excluded from the analysis. Genetic good risk ALL are defined by the presence of high hyperdiploidy, or *ETV6::RUNX1* fusion, or *DUX4/ERG*.

|  | Overall cohort |  |  | Genetic good risk ALL only |  |  |
| --- | --- | --- | --- | --- | --- | --- |
|  | *LZTR1*<sup>wt</sup>-ALL<br>N=1543 (%) | *LZTR1*<sup>mut/mut</sup>-ALL<br>N=19 (%) | p-value | *LZTR1*<sup>wt</sup>-ALL<br>N=704 (%) | *LZTR1*<sup>mut/mut</sup>-ALL<br>N=12 (%) | p-value |
| **Sex** |  |  |  |  |  |  |
| Male | 874 (56.6) | 7 (36.8) | ns | 371 (52.7) | 5 (41.7) | ns |
| Female | 669 (43.4) | 12 (63.2) |  | 333 (47.3) | 7 (58.3) |  |
| **Age** |  |  | ns |  |  | ns |
| <5 y | 718 (46.5) | 5 (26.3) |  | 381 (54.1) | 4 (33.3) |  |
| 5-9 y | 422 (27.3) | 9 (47.4) |  | 222 (31.5) | 5 (41.7) |  |
| ≥10 y | 403 (26.1) | 5 (26.3) |  | 101 (14.3) | 3 (25.0) |  |
| Median age (y) | 5.3 | 8 |  | 4.65 | 6.9 |  |
| **Immunophenotype** |  |  | ns |  |  | ns |
| T | 204 (13.2) | 1 (5.3) |  | 0 (0) | 0 (0.0) |  |
| B | 1339 (86.8) | 18 (94.7) |  | 704 (100) | 12 (100) |  |
| **WBC count (x10<sup>9</sup>/L)** |  |  | ns |  |  | ns |
| <10 | 480 (46.0) | 9 (50.0) |  | 301 (56.9) | 7 (63.6) |  |
| 10-<50 | 323 (30.9) | 6 (33.3) |  | 157 (29.7) | 2 (18.2) |  |
| 50-<100 | 112 (10.7) | 2 (11.1) |  | 55 (10.4) | 1 (9.1) |  |
| ≥100 | 129 (12.4) | 1 (5.6) |  | 16 (3.0) | 1 (9.1) |  |
| Missing | 499 | 1 |  | 175 | 1 |  |
| Median | 11.6 | 10.6 |  | 7.3 | 8.23 |  |
| **NCI risk group** |  |  | ns |  |  | ns |
| Standard risk | 596 (50.7) | 13 (68.4) |  | 382 (69.8) | 8 (66.7) |  |
| High risk | 580 (49.3) | 6 (31.6) |  | 165 (30.3) | 4 (33.3) |  |
| Missing | 367 | 0 |  | 157 | 0 |  |
| **CNS status** |  |  | ns |  |  | ns |
| CNS-1 | 762 (79.5) | 13 (92.9) |  | 428 (85.6) | 8 (100) |  |
| CNS-2 | 148 (15.4) | 0 (0.0) |  | 60 (12.0) | 0 (0.0) |  |
| CNS-3 | 49 (5.1) | 1 (7.1) |  | 12 (2.4) | 0 (0.0) |  |
| Missing | 584 | 5 |  | 204 | 4 |  |
| **Good Response to prephase** |  |  | ns |  |  | ns |
| No | 140 (12.4) | 2 (11.1) |  | 51 (8.9) | 1 (8.3) |  |
| Yes | 988 (87.6) | 16 (88.9) |  | 519 (91.1) | 11 (91.7) |  |
| Missing | 415 | 1 |  | 134 | 0 |  |
| **Genetic group** |  |  | 0.037 |  |  | ns |
| T | 204 (13.2) | 1 (5.3) |  | - | - |  |
| *ERG/DUX4* | 114 (8.5) | 1 (5.3) |  | 114 (16.2) | 1 (8.3) |  |
| *ETV6::RUNX1* | 248 (18.5) | 2 (10.5) |  | 248 (35.2) | 2 (16.7) |  |
| High hyperdiploidy | 342 (25.5) | 9 (47.4) |  | 342 (48.6) | 9 (75.0) |  |
| Low hypodiploidy | 6 (0.4) | 1 (5.3) |  | - | - |  |
| Near haploidy | 8 (0.6) | 1 (5.3) |  | - | - |  |
| *KMT2A-r* | 60 (4.5) | 0 (0.0) |  | - | - |  |
| *TCF3::PBX1* | 53 (4.0) | 0 (0.0) |  | - | - |  |
| *iAMP21* | 19 (1.4) | 0 (0.0) |  | - | - |  |
| Other | 489 (36.5) | 4 (21.1) |  | - | - |  |
| **Genetic risk group** |  |  | ns |  |  |  |
| Favorable risk | 704 (45.6) | 12 (63.2) |  | - | - |  |
| Unfavorable risk | 839 (54.4) | 7 (36.8) |  | - | - |  |
| **IKZF1del** | 221 (14.3) | 2 (10.5) | ns | 61 (8.7) | 1 (8.3) | ns |

wt: wild-type; mut: mutated; y: year; WBC: white blood cell; CNS: central nervous system ; ns : non-significant

**Table 3:**
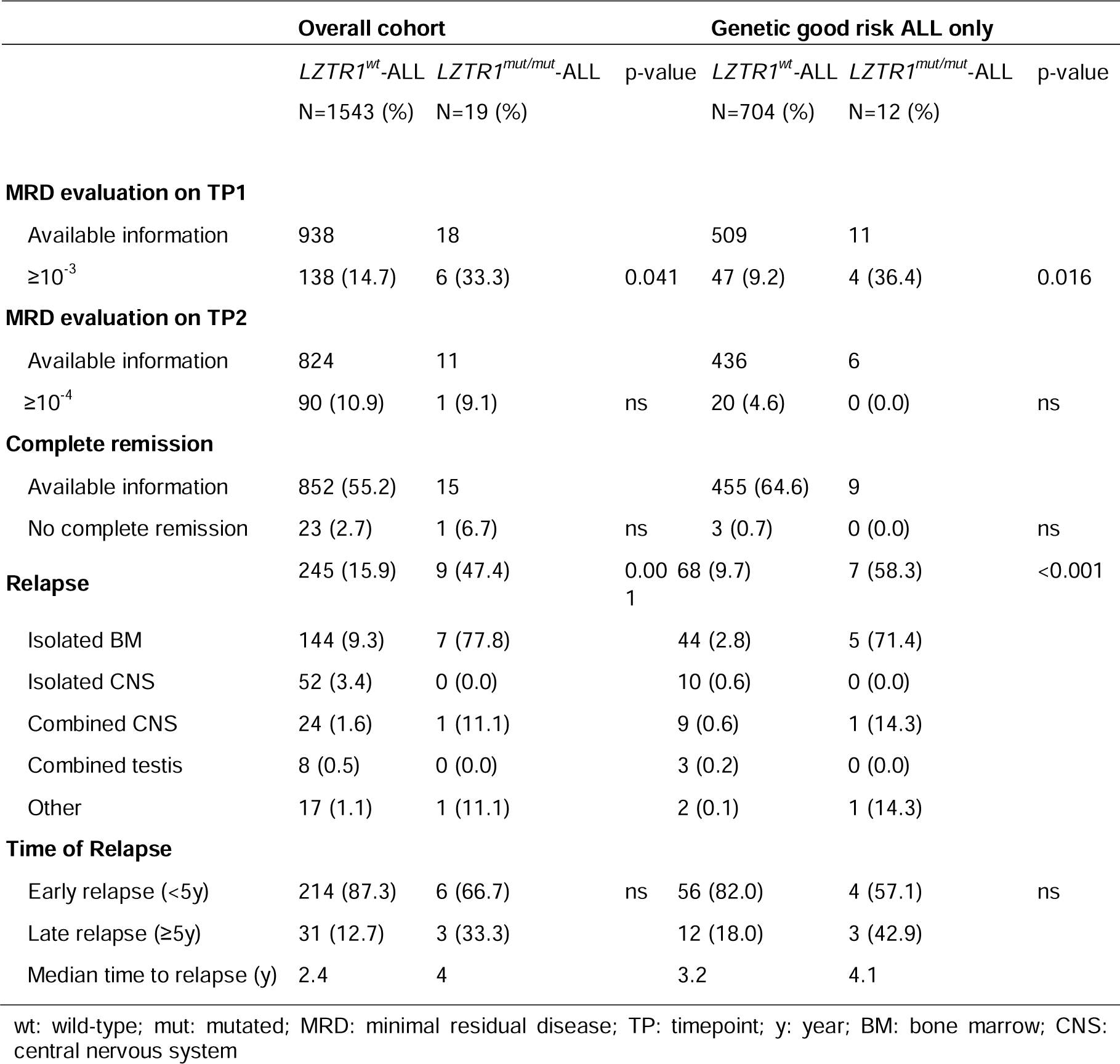
Treatment response and disease status according to the *LZTR1* status. . Patients with VUS (n=18) or monoallelic *LZTR1* variant (n=5) were excluded from the analysis. Genetic good risk ALL are defined by the presence of high hyperdiploidy, *ETV6::RUNX1* fusion, or *DUX4/ERG*. Results are expressed as number (%).

This was more pronounced in good-risk BCP-ALL: 7/12 (58.3%) patients with *LZTR1*^mut/mut^-ALL relapsed (4/11 HeH cases, 2/5 *ETV6::RUNX1*, 1/1 *ERG/DUX4* cases) versus 68/704 (9.7%) in *LZTR1^wt^*-ALL (p<0.001), leading to a 5-year EFS of 62.5% in *LZTR1*^mut/mut^-ALL versus 88.8% in *LZTR1^wt^*-ALL (HR: 5 [95% CI 2.3–10.85]; log-rank p<0.0001) (Figure 4B, right panel). Relapse rates were similar in patients with germline (5/14) and somatic (4/12) *LZTR1* alterations. Survival remained unaffected overall, though among 9 patients who relapsed, 1 with hypodiploid ALL died before reaching CR2, and 2 with HeH or *ETV6::RUNX1* ALL had a second relapse, one of whom died after a third relapse (Figure 2 and S8). *LZTR1* alterations thus define a subgroup with a high-risk of late-relapse within good-prognosis ALL.

### Expression of putative targets and functional inactivation of LZTR1

The identification of *LZTR1* as a driver in ALL prompted us to explore its expression pattern and that of its predicted substrates (H/N/KRAS, RIT1, AXL and EGFR)(11–13,30,31) across healthy hematopoietic compartments and in leukemia cell lines. *LZTR1* expression was low but ubiquitous across hematopoietic stem/progenitor cells (HSPCs)(32), single-cell bone marrow profiles (n=146)(33), and cell lines, without variation by lineage or ontogeny (Figure S9, S10).

In contrast, LZTR1 targets displayed lineage specificity. Canonical RAS isoforms (H/K/NRAS) were uniformly expressed, whereas non-canonical MRAS, RIT1 and AXL peaked in the granulocyte-monocyte lineage (Figure S9). Protein analysis confirmed that MRAS, RIT1 and AXL were largely restricted to myeloid lines (Figure S10). EGFR was undetectable in hematopoietic cells.

To assess the functional consequences of *LZTR1* loss, we generated *LZTR1*-knockout cell lines using CRISPR–Cas9 editing. Bi-allelic *LZTR1* knockout increased canonical RAS protein levels (Figures 5A, S11A). RIT1 showed strong upregulation across both lymphoid and myeloid lineages. MRAS accumulated only in myeloid cells. Heterozygous *LZTR1*^⁺/⁻^ clones showed no detectable changes, indicating that bi-allelic alteration is required (Figure S12).

**Figure 5:**
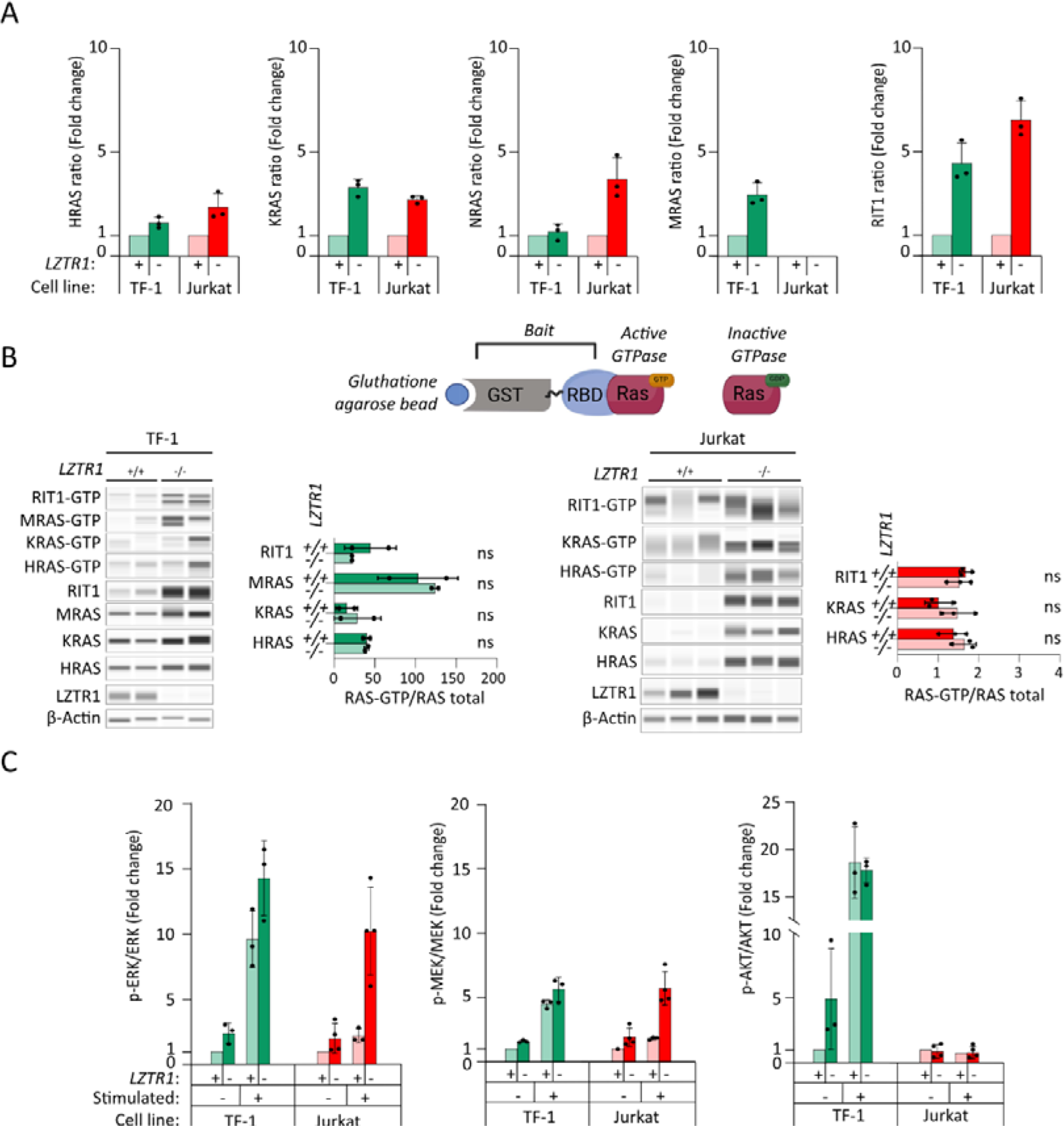
Consequences of functional inactivation of *LZTR1*. **A.** Effect of *LZTR1* knock-out on RAS protein expression in TF-1 and Jurkat cell lines (expressed as fold-change relative to the wild-type used as baseline and set to 1. Dots indicate individual replicates. **B.** Schematic of the GST–RAS-binding domain (RBD) pull-down which captures active, GTP-bound, small GTPases (upper panel); Representative immunoblots and quantification for HRAS-GTP, KRAS-GTP, NRAS-GTP, and MRAS-GTP after stimulation by granulocyte–macrophage colony-stimulating factor (GM-CSF) (TF-1) or phorbol-12-myristate-13-acetate (PMA)-ionomycin (Jurkat). Total RAS isoform levels are shown in parallel. For each isoform, the GTP-bound signal was normalized to the total input of that isoform. **C.** Downstream signaling readouts pMEK/MEK, pERK/ERK, and pAKT/AKT ratios in TF-1 and Jurkat cell lines, at baseline and after stimulation (expressed as fold-change relative to the wild-type used as baseline and set to 1, with dots indicating replicate measurements.

Pull-down assays showed that RAS-GTP/GDP ratios were unaltered in *LZTR1*^-/-^ clones versus *LZTR1*^+/+^, even upon stimulation of the RAS pathway, indicating that LZTR1 regulates total RAS pool abundance rather than GTP-bound forms (Figure 5B). *LZTR1*^-/-^ cells showed modest phospho-ERK/MEK elevation without phospho-AKT changes (Figure 5C and S11B).

These data demonstrate that LZTR1 controls steady-state RAS protein levels independent of activation status and specifically represses non-canonical RIT1 in lymphoid cells.

### LZTR1 and targets in BCP-ALL

We next examined LZTR1 expression in primary ALL samples. *LZTR1* transcript levels were significantly higher in *LZTR1^wt^-*BCP-ALL diagnosis samples than in matched remission samples or *LZTR1^mut/mut^*samples (Figure 6A). The difference was more pronounced at the protein level (Figure 6BE). LZTR1 expression positively correlated with RAS-activating mutations (R²=0.39; p=0.0059; Figure 6C), suggesting compensatory upregulation as negative feedback to oncogenic RAS signaling.

**Figure 6:**
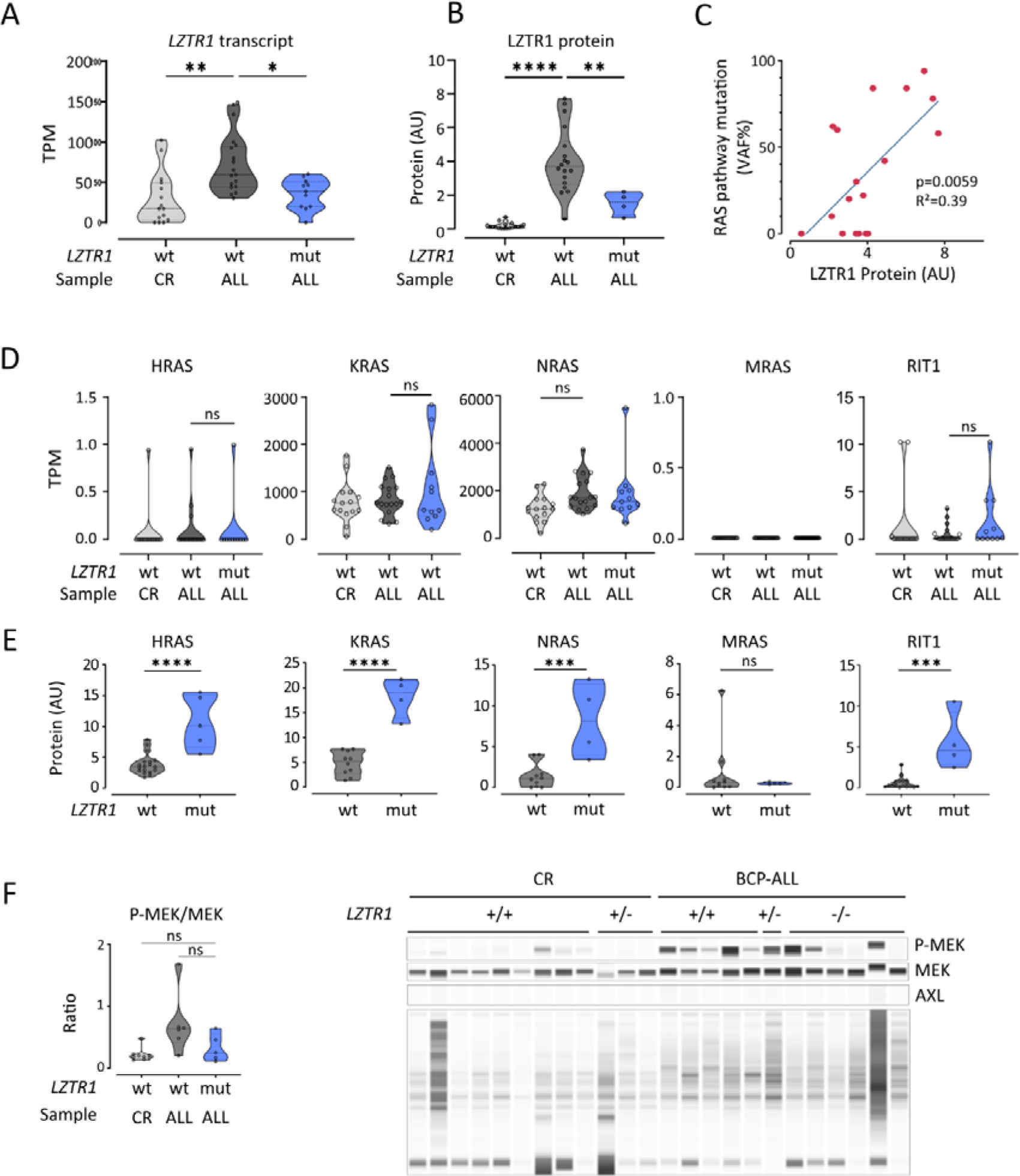
Effect of *LZTR1* bi-allelic mutation in BCP-ALL. **A.** *LZTR1* transcript expression at diagnosis of *LZTR1* wild-type (*LZTR1*^wt^-ALL; n=17) or mutated (*LZTR1*^mut/mut^-ALL; n=11) BCP-ALL, and at complete remission (*LZTR1*^wt^-CR; n=15). Expression levels were expressed in transcripts per million (TPM). **B.** LZTR1 protein expression at diagnosis of *LZTR1* wild-type (*LZTR1*^wt^-CR; n=18) or bi-allelic mutated (mut; n=4) BCP-ALL, and at complete remission (n=15). LZTR1 protein are normalized on total protein and expressed in arbitrary units (AU). **C**. Correlation between the amount of LZTR1 protein and the fraction of RAS-pathway-mutated leukemia in *LZTR1*^wt^-BCP-ALL (n=18); P value and R² are indicated. **D.** Transcript expression of RAS family members (*HRAS*, *KRAS*, *NRAS*, *MRAS* and *RIT1*) at diagnosis in *LZTR1*^wt^-ALL (n = 17) or *LZTR1*^mut/mut^-BCP-ALL (n = 11), and at complete remission (n=15). Expression levels were quantified by RNA-seq and expressed in transcripts per million (TPM). **E.** RAS-family (HRAS, KRAS, NRAS, MRAS and RIT1) protein expression at diagnosis of *LZTR1*^wt^-ALL (n=10) or bi-allelic *LZTR1*^mut/mut^-ALL (n=4) BCP-ALL. Protein expression is normalized on total protein. **F.** MAPK pathway assessed by measuring the phospho-MEK/MEK protein ratio in *LZTR1*^wt^-ALL (n=6) or bi-allelic *LZTR1*^mut/mut^-BCP-ALL (n=5), and at complete remission (n=9). Protein expression is normalized on total protein.

All *RAS* genes (except *MRAS*) were transcribed across BCP-ALL genetic subtypes, whereas *AXL* and *EGFR* transcripts were largely undetectable (Figure S13). Transcript levels of LZTR1 targets showed no significant differences between diagnostic and CR samples, nor between *LZTR1*^mut/mut^-ALL and *LZTR1*^wt^-ALL cases (Figure 6D and S13). In contrast, canonical RAS protein levels were significantly higher in *LZTR1*^mut/mut^-ALL compared to *LZTR1*^wt^-ALL. Most strikingly, RIT1 protein, normally absent from healthy lymphoid cells and *LZTR1*^wt^-ALL, was strongly expressed in *LZTR1*^mut/mut^-ALL. MRAS and EGFR remained undetectable (Figure 6E and S10). MAPK activation was not significantly increased in *LZTR1*^mut/mut^-ALL (Figure 6F). These observations reveal that *LZTR1* inactivation in ALL drives accumulation of RAS-family proteins, including ectopic lymphoid expression of RIT1.

### Drug response profiling

Drug response profiling was performed on 4 *LZTR1*^mut*/*mut^-ALL PDX samples at diagnosis (#D-23031 and #D-17639) and relapse (#R-28839 and #R-22646) (Figure 7). *LZTR1*^mut*/*mut^-ALL showed a response to standard induction drugs comparable to other ALL. Relapse samples exhibited greater resistance. Diagnosis samples were sensitive to MEK inhibitors (trametinib, selumetinib), whereas relapse samples were resistant.

**Figure 7:**
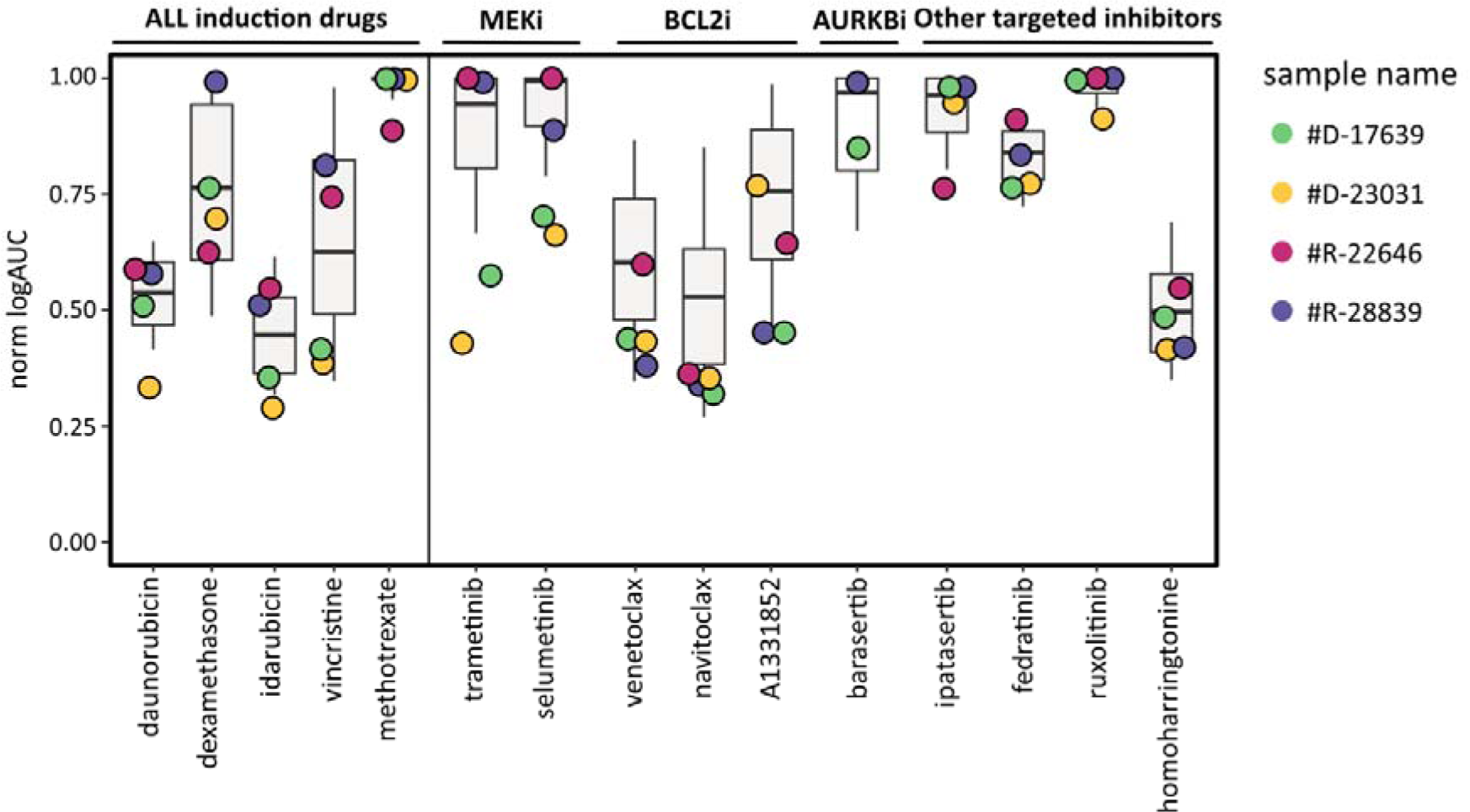
Drug sensitivity pattern of ALL with bi-allelic *LZTR1* alterations. *Ex Vivo* drug response profiling of 4 *LZTR1^mut/mut^*-ALL PDX samples is represented as normalized logAUC for indicated compounds (n=4 samples; A1331852, venetoclax, navitoclax and barasertib screened in n=2 samples). Data are referenced against a general ALL patient cohort (boxplot)(27). AURKBi: Aurora Kinase B inhibitor.

Aurora B inhibitor barasertib (#D-17639, #R-28839) showed no response, despite RIT1 reported role in spindle assembly checkpoint impairment and aneuploidy(34,35). Notably, *LZTR1*^mut/mut^-ALL demonstrated high sensitivity to BCL2 family inhibitors (venetoclax, navitoclax).

## Discussion

Previous reports suggested that *LZTR1* alterations might be associated with an increased risk of leukemia(16,17,19). Here, screening a large cohort of children with ALL revealed somatic and/or germline pathogenic *LZTR1* variants in 1.6% of patients. *LZTR1* variants were enriched in HeH ALL but were also found in other genetic subgroups of ALL, unlike what has been reported for *PTPN11* mutations(10).

LZTR1 shows weak evolutionary constraint with 0.31% truncating variant prevalence in gnomAD (1/320)(36). Using schwannomatosis methodology(22), we estimate ∼0.1% penetrance for ALL development in childhood(37) (1/316,000), mirroring low-penetrance schwannomatosis and excluding major monogenic predisposition status. This informs germline screening interpretation, as *LZTR1* variants may be incidental (38).

LZTR1 acts as a classical tumour suppressor gene in ALL, following a two-hit model, in which germline or somatic loss of function in one *LZTR1* allele contributes to leukemogenesis upon inactivation of the second allele through point mutations, 22qCN-LOH, or RNA LOH. The loss of a single *LZTR1* allele is insufficient to induce biological changes and activate RIT1 expression. In support of this model, mouse *Lztr1*-KO studies showed that half of mice reconstituted with *Lztr1*-KO fetal liver cells developed B-cell ALL(39).

Recurrent 22qCN-LOH has not been reported so far in BCP-ALL(40), and strictly co-segregated with *LZTR1* variants, absent in controls. Notably, CN-LOH was never associated with truncating mutations, and the preference for CN-LOH or RNA LOH over deletions supports the requirement for residual LZTR1 activity for cell viability, extending this observation to hematopoietic cells(13,17,39,41).

Why CN-LOH or RNA-LOH is favored over gene deletion to inactivate the second allele remains unclear. No alternative 22q targets were identified in ALL. A possibility is that the preservation of surrounding gene expression is under strong selective pressure in leukemia cells. In line with this hypothesis, the lower schwannomatosis risk recently reported in patients with DiGeorge 22q11.2 deletions supports gene dosage constraints(42).

*LZTR1*^mut/mut^-ALL lacked defining transcriptional signatures but showed induction resistance, slow MRD clearance, and high late relapse risk (5-year EFS 62.5% vs 88.8%; HR 5.0) within good-prognosis subtypes, challenging de-escalation strategies. (43,44)

LZTR1 upregulation in *LZTR1^wt^*-ALL correlated with RAS activating mutations, indicating compensatory feedback. Bi-allelic loss abolished this regulation and drove canonical RAS accumulation and ectopic lymphoid RIT1 expression, normally myeloid-restricted. RIT1 detection in lymphoid cells can thus be used as an effective functional test to assess the pathogenicity of *LZTR1* variants.

*RIT1* is rarely mutated in ALL(45), and mice with RIT1 activation develop MPN but no B-cell ALL(39), suggesting the transforming cellular effects of LZTR1 loss on lymphoid cells are not entirely reliant on RIT1. RIT1 shares activation of the MAPK pathway with other Ras GTPases. This aligns with the sensitivity to MEK-inhibitors observed in the *LZTR1*^mut/mut^ samples tested at diagnosis. Lack of sensitivity in the two relapse samples indicates that LZTR1 loss may not be the only determinant of sensitivity to MEK inhibitors. Preliminary evidence of sensitivity to BCL2 inhibitors warrants further investigation and supports the potential of functional drug profiling to identify actionable vulnerabilities.

In conclusion, LZTR1 does not qualify as a high-penetrance Mendelian predisposition gene but functions as a *bona fide* tumor suppressor in ALL leukemogenesis through a classical two-hit mechanism. Bi-allelic *LZTR1* alterations identify patients at high risk of late relapse within otherwise favorable-risk genetic subgroups, informing against treatment de-escalation. The mechanisms underlying the oncogenic effects of LZTR1 and RIT1 are still elusive, but elucidating them will be important for guiding treatment.

## Abbreviations

AD: Autosomal dominant
ALL: Acute lymphoblastic leukemia
AR: Autosomal recessive
BAF: B-allele frequency
BCP-ALL: B-cell precursor acute lymphoblastic leukemia
BM: Bone marrow
CIR: Cumulative incidence of relapse
CNA: Copy-number alteration
CN-LOH: Copy-neutral loss of heterozygosity
CR: Complete remission
DN: Dominant negative
DRP: Drug response profiling
EFS: Event-free survival
FBS: Fetal bovine serum
GM-CSF: Granulocyte–macrophage colony-stimulating factor
HeH: High hyperdiploidy
HGVS: Human Genome Variation Society
HR: Hazard ratio
HSPC: Hematopoietic stem/progenitor cell
IRB: Institutional review board
LoF: Loss of function
*LZTR1*^mut^-ALL: Acute lymphoblastic leukemia with LZTR1-alteration
*LZTR1*^wt^-ALL: Acute lymphoblastic leukemia without
LZTR1 alteration *LZTR1*^mut/mut^-ALL: Acute lymphoblastic leukemia with bi-allelic
LZTR1 alteration MAPK: Mitogen-activated protein kinase
MLPA: Multiplex ligation-dependent probe amplification
MRD: Minimal residual disease
NGS: Next-generation sequencing
NS: Noonan syndrome
OS: Overall survival
P/LP: Pathogenic/likely pathogenic
PAM: Protospacer adjacent motif
PCA: Principal component analysis
PDX: Patient-derived xenograft
PMA: Phorbol-12-myristate-13-acetate
RNA LOH: RNA-level loss of heterozygosity
RNA-seq: RNA sequencing
SNP: Single-nucleotide polymorphism
VAF: Variant allele frequency
VUS: Variant of uncertain significance
WES: Whole-exome sequencing
wt: Wild-type

## Data Availability

The data that support the findings of this study are available from the corresponding author upon reasonable request. RNA sequencing data from healthy hematopoietic samples are publicly available in the Gene Expression Omnibus (GEO) repository under accession number GSE267628 (https://www.ncbi.nlm.nih.gov/geo/query/acc.cgi?acc=GSE267628)

## Acknowledgments

We thank patients’ families and pediatric oncologists from the “Société Française de lutte contre les cancers et leucémies de l’enfant et de l’adolescent” (SFCE) for their participation to the research. We thank Prof. Sylvie Chevret for reviewing statistical analyses. We thank Thomas Mercher (INSERM, Institut Gustave Roussy, Villejuif, France) for insightful discussion and review of the manuscript. We thank Aldjia Assous for help in data management. We thank the Center for Biological Resources (CRB-cancer) (BB-0033-00076) of the Robert Debré hospital.

## Conflicts of Interest

The authors declare no conflicts of interest.

## Funding

The authors declare that this research did not receive funding.

## Notes

### Competing Interest Statement

The authors have declared no competing interest.

### Author Declarations

The Institutional Review Board of Hopitaux Universitaires Paris-Nord Val-de-Seine gave ethical approval for this work (IRB-00006477). The study was conducted in accordance with the Declaration of Helsinki.

## References

1. Jeha S, Choi J, Roberts KG, Pei D, Coustan-Smith E, Inaba H, et al. Clinical significance of novel subtypes of acute lymphoblastic leukemia in the context of minimal residual disease-directed therapy. Blood Cancer Discov. 2021 Jul;2(4):326–37. doi:10.1158/2643-3230.BCD-20-0229 PubMed PMID: 34250504; PubMed Central PMCID: PMC8265990.

2. Clappier E, Auclerc MF, Rapion J, Bakkus M, Caye A, Khemiri A, et al. An intragenic ERG deletion is a marker of an oncogenic subtype of B-cell precursor acute lymphoblastic leukemia with a favorable outcome despite frequent IKZF1 deletions. Leukemia. 2014 Jan;28(1):70–7. doi:10.1038/leu.2013.277 PubMed PMID: 24064621.

3. Mattano LA, Devidas M, Maloney KW, Wang C, Friedmann AM, Buckley P, et al. Favorable Trisomies and ETV6-RUNX1 Predict Cure in Low-Risk B-Cell Acute Lymphoblastic Leukemia: Results From Children’s Oncology Group Trial AALL0331. J Clin Oncol. 2021 May 10;39(14):1540–52. doi:10.1200/JCO.20.02370 PubMed PMID: 33739852; PubMed Central PMCID: PMC8274747.

4. Purvis K, Zhou Y, Karol SE, Rubnitz JE, Ribeiro RC, Lee S, et al. Outcomes in patients with ETV6::RUNX1 or high-hyperdiploid B-ALL treated in the St. Jude Total Therapy XV/XVI studies. Blood. 2025 Jan 9;145(2):190–201. doi:10.1182/blood.2024024936 PubMed PMID: 39316653; PubMed Central PMCID: PMC11738036.

5. Schore RJ, Angiolillo AL, Kairalla JA, Devidas M, Rabin KR, Zweidler-McKay P, et al. Outstanding outcomes with two low intensity regimens in children with low-risk B-ALL: a report from COG AALL0932. Leukemia. 2023 Jun;37(6):1375–8. doi:10.1038/s41375-023-01870-8 PubMed PMID: 36966262; PubMed Central PMCID: PMC10503688.

6. Enshaei A, Vora A, Harrison CJ, Moppett J, Moorman AV. Defining low-risk high hyperdiploidy in patients with paediatric acute lymphoblastic leukaemia: a retrospective analysis of data from the UKALL97/99 and UKALL2003 clinical trials. Lancet Haematol. 2021 Nov;8(11):e828–39. doi:10.1016/S2352-3026(21)00304-5 PubMed PMID: 34715050; PubMed Central PMCID: PMC8567211.

7. Dastugue N, Suciu S, Plat G, Speleman F, Cavé H, Girard S, et al. Hyperdiploidy with 58-66 chromosomes in childhood B-acute lymphoblastic leukemia is highly curable: 58951 CLG-EORTC results. Blood. 2013 Mar 28;121(13):2415–23. doi:10.1182/blood-2012-06-437681 PubMed PMID: 23321258.

8. Piette C, Suciu S, Clappier E, Bertrand Y, Drunat S, Girard S, et al. Differential impact of drugs on the outcome of ETV6-RUNX1 positive childhood B-cell precursor acute lymphoblastic leukaemia: results of the EORTC CLG 58881 and 58951 trials. Leukemia. 2018 Jan;32(1):244–8. doi:10.1038/leu.2017.289 PubMed PMID: 29064485.

9. Klco JM, Mullighan CG. Advances in germline predisposition to acute leukaemias and myeloid neoplasms. Nat Rev Cancer. 2021 Feb;21(2):122–37. doi:10.1038/s41568-020-00315-z PubMed PMID: 33328584; PubMed Central PMCID: PMC8404376.

10. Cavé H, Caye A, Strullu M, Aladjidi N, Vignal C, Ferster A, et al. Acute lymphoblastic leukemia in the context of RASopathies. European Journal of Medical Genetics. 2016 Mar;59(3):173–8. doi:10.1016/j.ejmg.2016.01.003

11. Bigenzahn JW, Collu GM, Kartnig F, Pieraks M, Vladimer GI, Heinz LX, et al. LZTR1 is a regulator of RAS ubiquitination and signaling. Science. 2018 Dec 7;362(6419):1171–7. doi:10.1126/science.aap8210

12. Castel P, Cheng A, Cuevas-Navarro A, Everman DB, Papageorge AG, Simanshu DK, et al. RIT1 oncoproteins escape LZTR1-mediated proteolysis. Science. 2019 Mar 15;363(6432):1226–30. doi:10.1126/science.aav1444

13. Steklov M, Pandolfi S, Baietti MF, Batiuk A, Carai P, Najm P, et al. Mutations in LZTR1 drive human disease by dysregulating RAS ubiquitination. Science. 2018 Dec 7;362(6419):1177–82. doi:10.1126/science.aap7607

14. Frattini V, Trifonov V, Chan JM, Castano A, Lia M, Abate F, et al. The integrated landscape of driver genomic alterations in glioblastoma. Nat Genet. 2013 Oct;45(10):1141–9. doi:10.1038/ng.2734 PubMed PMID: 23917401; PubMed Central PMCID: PMC3799953.

15. Piotrowski A, Xie J, Liu YF, Poplawski AB, Gomes AR, Madanecki P, et al. Germline loss-of-function mutations in LZTR1 predispose to an inherited disorder of multiple schwannomas. Nat Genet. 2014 Feb;46(2):182–7. doi:10.1038/ng.2855 PubMed PMID: 24362817; PubMed Central PMCID: PMC4352302.

16. Yamamoto GL, Aguena M, Gos M, Hung C, Pilch J, Fahiminiya S, et al. Rare variants in SOS2 and LZTR1 are associated with Noonan syndrome. J Med Genet. 2015 Jun;52(6):413–21. doi:10.1136/jmedgenet-2015-103018

17. Johnston JJ, van der Smagt JJ, Rosenfeld JA, Pagnamenta AT, Alswaid A, Baker EH, et al. Autosomal recessive Noonan syndrome associated with biallelic LZTR1 variants. Genet Med. 2018;20(10):1175–85. doi:10.1038/gim.2017.249 PubMed PMID: 29469822; PubMed Central PMCID: PMC6105555.

18. Chinton J, Huckstadt V, Mucciolo M, Lepri F, Novelli A, Gravina LP, et al. Providing more evidence on LZTR1 variants in Noonan syndrome patients. Am J Med Genet. 2020 Feb;182(2):409–14. doi:10.1002/ajmg.a.61445

19. Unuma K, Tomomasa D, Noma K, Yamamoto K, Matsuyama T aki, Makino Y, et al. Case Report: Molecular autopsy underlie COVID-19-associated sudden, unexplained child mortality. Front Immunol. 2023 Apr 18;14:1121059. doi:10.3389/fimmu.2023.1121059 PubMed PMID: 37143668; PubMed Central PMCID: PMC10151512.

20. Zipper L, Wagener R, Fischer U, Hoffmann A, Yasin L, Brandes D, et al. Hyperdiploid acute lymphoblastic leukemia in children with *LZTR1* germline variants. HemaSphere. 2024 Jan;8(1). doi:10.1002/hem3.26

21. Karczewski KJ, Francioli LC, Tiao G, Cummings BB, Alföldi J, Wang Q, et al. The mutational constraint spectrum quantified from variation in 141,456 humans. Nature. 2020 May;581(7809):434–43. doi:10.1038/s41586-020-2308-7

22. Deng F, Evans DG, Smith MJ. Comparison of the frequency of loss-of-function LZTR1 variants between schwannomatosis patients and the general population. Hum Mutat. 2022 Jul;43(7):919–27. doi:10.1002/humu.24376 PubMed PMID: 35391499; PubMed Central PMCID: PMC9324957.

23. Li MM, Datto M, Duncavage EJ, Kulkarni S, Lindeman NI, Roy S, et al. Standards and Guidelines for the Interpretation and Reporting of Sequence Variants in Cancer: A Joint Consensus Recommendation of the Association for Molecular Pathology, American Society of Clinical Oncology, and College of American Pathologists. J Mol Diagn. 2017 Jan;19(1):4–23. doi:10.1016/j.jmoldx.2016.10.002 PubMed PMID: 27993330; PubMed Central PMCID: PMC5707196.

24. Richards S, Aziz N, Bale S, Bick D, Das S, Gastier-Foster J, et al. Standards and guidelines for the interpretation of sequence variants: a joint consensus recommendation of the American College of Medical Genetics and Genomics and the Association for Molecular Pathology. Genetics in Medicine. 2015 May;17(5):5. doi:10.1038/gim.2015.30

25. Schmitz M, Breithaupt P, Scheidegger N, Cario G, Bonapace L, Meissner B, et al. Xenografts of highly resistant leukemia recapitulate the clonal composition of the leukemogenic compartment. Blood. 2011 Aug 18;118(7):1854–64. doi:10.1182/blood-2010-11-320309 PubMed PMID: 21670474.

26. Frismantas V, Dobay MP, Rinaldi A, Tchinda J, Dunn SH, Kunz J, et al. Ex vivo drug response profiling detects recurrent sensitivity patterns in drug-resistant acute lymphoblastic leukemia. Blood. 2017 Mar 16;129(11):e26–37. doi:10.1182/blood-2016-09-738070

27. Steffen FD, Lissat A, Alten J, Eckert C, Bodmer N, Thorhauge Als-Nielsen BE, et al. Drug Response Profiling Informs Personalized Bridging to Cell Therapy for Patients with Relapsed/Refractory Acute Lymphoblastic Leukemia. Blood. 2023 Nov 2;142(Supplement 1):4350. doi:10.1182/blood-2023-179306

28. Ritz C, Baty F, Streibig JC, Gerhard D. Dose-Response Analysis Using R. PLOS ONE. 2015 Dec 30;10(12):e0146021. doi:10.1371/journal.pone.0146021

29. Li Q, Wang K. InterVar: Clinical Interpretation of Genetic Variants by the 2015 ACMG-AMP Guidelines. The American Journal of Human Genetics. 2017 Feb;100(2):267–80. doi:10.1016/j.ajhg.2017.01.004

30. Ko A, Hasanain M, Oh YT, D’Angelo F, Sommer D, Frangaj B, et al. LZTR1 Mutation Mediates Oncogenesis through Stabilization of EGFR and AXL. Cancer Discov. 2023 Mar 1;13(3):702–23. doi:10.1158/2159-8290.CD-22-0376 PubMed PMID: 36445254.

31. Motta M, Fidan M, Bellacchio E, Pantaleoni F, Schneider-Heieck K, Coppola S, et al. Dominant Noonan syndrome-causing LZTR1 mutations specifically affect the Kelch domain substrate-recognition surface and enhance RAS-MAPK signaling. Human Molecular Genetics. 2019 Mar 15;1007–22.

32. Strullu M, Arfeuille C, Caye-Eude A, Maillard L, Lainey E, Piques F, et al. Two distinct fetal-type signatures characterize juvenile myelomonocytic leukemia. Hemasphere. 2025 Mar;9(3):e70104. doi:10.1002/hem3.70104 PubMed PMID: 40134525; PubMed Central PMCID: PMC11934893.

33. Li M, Zhang X, Ang KS, Ling J, Sethi R, Lee NYS, et al. DISCO: a database of Deeply Integrated human Single-Cell Omics data. Nucleic Acids Res. 2022 Jan 7;50(D1):D596–602. doi:10.1093/nar/gkab1020 PubMed PMID: 34791375; PubMed Central PMCID: PMC8728243.

34. Cuevas-Navarro A, Van R, Cheng A, Urisman A, Castel P, McCormick F. The RAS GTPase RIT1 compromises mitotic fidelity through spindle assembly checkpoint suppression. Current Biology. 2021 Sep;31(17):3915–3924.e9. doi:10.1016/j.cub.2021.06.030

35. Vichas A, Riley AK, Nkinsi NT, Kamlapurkar S, Parrish PCR, Lo A, et al. Integrative oncogene-dependency mapping identifies RIT1 vulnerabilities and synergies in lung cancer. Nat Commun. 2021 Aug 9;12(1):4789. doi:10.1038/s41467-021-24841-y PubMed PMID: 34373451; PubMed Central PMCID: PMC8352964.

36. Stoltze UK, Foss-Skiftesvik J, Hansen TVO, Rasmussen S, Karczewski KJ, Wadt KAW, et al. The evolutionary impact of childhood cancer on the human gene pool. Nat Commun. 2024 Feb 29;15(1):1881. doi:10.1038/s41467-024-45975-9

37. Malard F, Mohty M. Acute lymphoblastic leukaemia. Lancet. 2020 Apr 4;395(10230):1146–62. doi:10.1016/S0140-6736(19)33018-1 PubMed PMID: 32247396.

38. Perrino MR, Das A, Scollon SR, Mitchell SG, Greer MLC, Yohe ME, et al. Update on Pediatric Cancer Surveillance Recommendations for Patients with Neurofibromatosis Type 1, Noonan Syndrome, CBL Syndrome, Costello Syndrome, and Related RASopathies. Clinical Cancer Research. 2024 Nov 1;30(21):4834–43. doi:10.1158/1078-0432.CCR-24-1611

39. Chen S, Vedula RS, Cuevas-Navarro A, Lu B, Hogg SJ, Wang E, et al. Impaired Proteolysis of Noncanonical RAS Proteins Drives Clonal Hematopoietic Transformation. Cancer Discov. 2022 Oct 5;12(10):2434–53. doi:10.1158/2159-8290.CD-21-1631 PubMed PMID: 35904492; PubMed Central PMCID: PMC9533010.

40. Lundin-Ström KB, Biloglav A, Lilljebjörn H, Rissler M, Fioretos T, Hansson M, et al. Whole-exome sequencing exploration of acquired uniparental disomies in B-cell precursor acute lymphoblastic leukemia. Leukemia. 2018 Sep;32(9):2058–62. doi:10.1038/s41375-018-0191-0

41. Shamseldin HE, Kurdi W, Almusafri F, Alnemer M, Alkaff A, Babay Z, et al. Molecular autopsy in maternal–fetal medicine. Genetics in Medicine. 2018 Apr;20(4):420–7. doi:10.1038/gim.2017.111

42. Evans DG, Messiaen LM, Foulkes WD, Irving REA, Murray AJ, Perez-Becerril C, et al. Typical 22q11.2 deletion syndrome appears to confer a reduced risk of schwannoma. Genetics in Medicine. 2021 Sep;23(9):1779–82. doi:10.1038/s41436-021-01175-0

43. Domenech C, Kicinski M, De Moerloose B, Piette C, Chahla WA, Kornreich L, et al. Results of the prospective EORTC Children Leukemia Group study 58081 in precursor B- and T-cell acute lymphoblastic leukemia. Hemasphere. 2024 Nov 13;8(11):e70025. doi:10.1002/hem3.70025 PubMed PMID: 39540141; PubMed Central PMCID: PMC11558101.

44. Chang TC, Chen W, Qu C, Cheng Z, Hedges D, Elsayed A, et al. Genomic Determinants of Outcome in Acute Lymphoblastic Leukemia. J Clin Oncol. 2024 Oct 10;42(29):3491–503. doi:10.1200/JCO.23.02238 PubMed PMID: 39121442; PubMed Central PMCID: PMC11458106.

45. Cavé H, Caye A, Ghedira N, Capri Y, Pouvreau N, Fillot N, et al. Mutations in RIT1 cause Noonan syndrome with possible juvenile myelomonocytic leukemia but are not involved in acute lymphoblastic leukemia. Eur J Hum Genet. 2016 Aug;24(8):1124–31. doi:10.1038/ejhg.2015.273 PubMed PMID: 26757980; PubMed Central PMCID: PMC4970687.

