## Supplemental figures 1-13 for "LZTR1 functions as a two-hit tumor suppressor in childhood acute lymphoblastic leukemia"

**Supplementary figures**

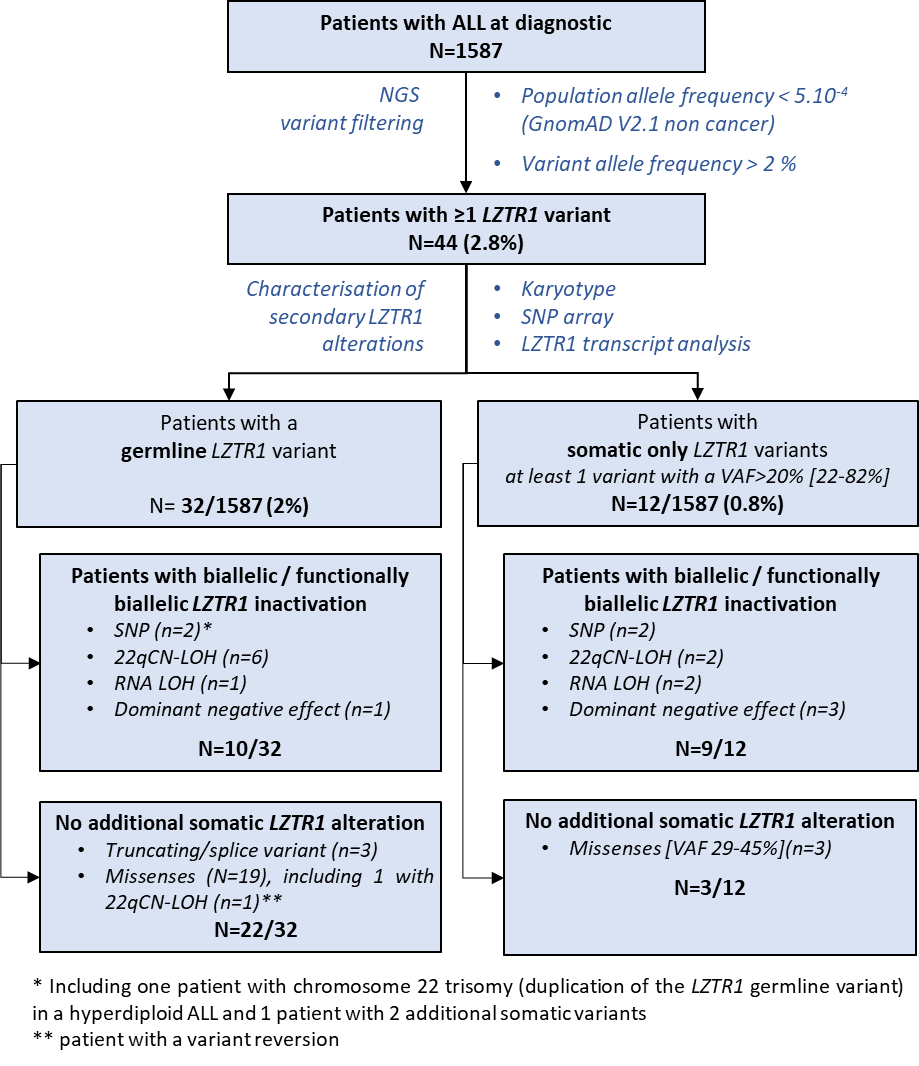
**Figure S1** **: Patients flowchart.** NGS: Next generation sequencing; VAF: variant allele frequency; CN-LOH: copy number neutral-loss of heterozygosity; P/LP : probably pathogenic/pathogenic; VUS: variant of unknown significance

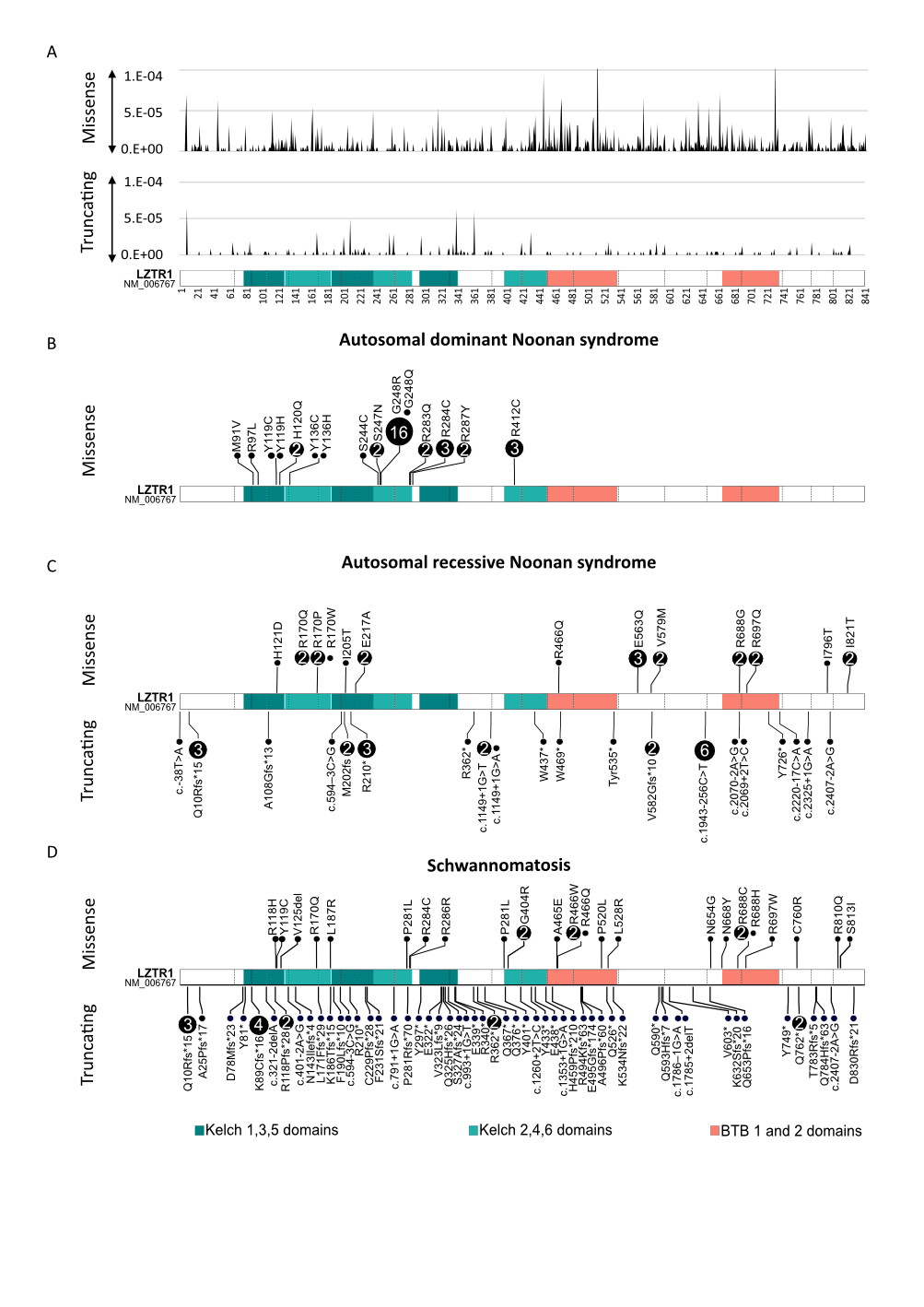
**Figure S2. Distribution of *LZTR1* variants in Noonan syndrome and schwannomatosis. A.** Frequency of missense (top) and truncating/splice (bottom) variants in gnomAD v2.1.1, mapped along the protein (Kelch 1–6 and BTB1–2 domains). Missense variants are enriched toward the C-terminal region, whereas truncating variants are distributed across the gene. In gnomAD v2, the overall frequency of non-synonymous missense variants ranges from 0.74% in Europeans to 2.5% in African/African American populations. The frequency of heterozygous truncating variants ranges from 0.27% in Ashkenazi Jewish individuals to 0.5% in African/African American populations. **B-D.** Distribution of pathogenic or likely pathogenic variants in autosomal dominant Noonan syndrome (B), autosomal recessive Noonan syndrome (C), and schwannomatosis (D). Variants in AD Noonan syndrome are mainly missense and cluster within Kelch domains. Shared variants or affected residues across conditions suggest common mutational hotspots.

**Figure S3: Functional analyses of variants of unknown significance in *LZTR1.* A*.*** Aberrant splicing in hypodiploid ALL. Sashimi plots from bulk RNA-seq (read coverage and exon–exon junctions) reveal cryptic splice-site usage. Left (Pt #24596): creation of a novel 3′ splice site within intron 2 inserts the final 11 nucleotides of intron 2, producing the frameshift p.Lys86Cysfs*16. Right (Pt #28956): activation of an alternative 5′ splice site within exon 17 deletes the first 17 nucleotides of exon 17, yielding p.Thr649Hisfs*14. **B.** Pathogenicity assessment of the c.2248_2256delGGCTTCTAC (p.Gly750_Tyr752del) in-frame deletion. The variant is absent from control population databases and and removes a highly conserved glycine residue (conserved across species to *D. melanogaster*Structural modeling predicts a change in the BTB-domain dimerization/CUL3-binding angle from ~19 Å to ~9 Å in the critical region necessary for LZTR1 dimerization and CUL3 binding. Consistent with its pathogenicity, RNA-seq detected the 9-nucleotide deletion in more than 50% of the reads within the generated BAM files (RNA-LOH).

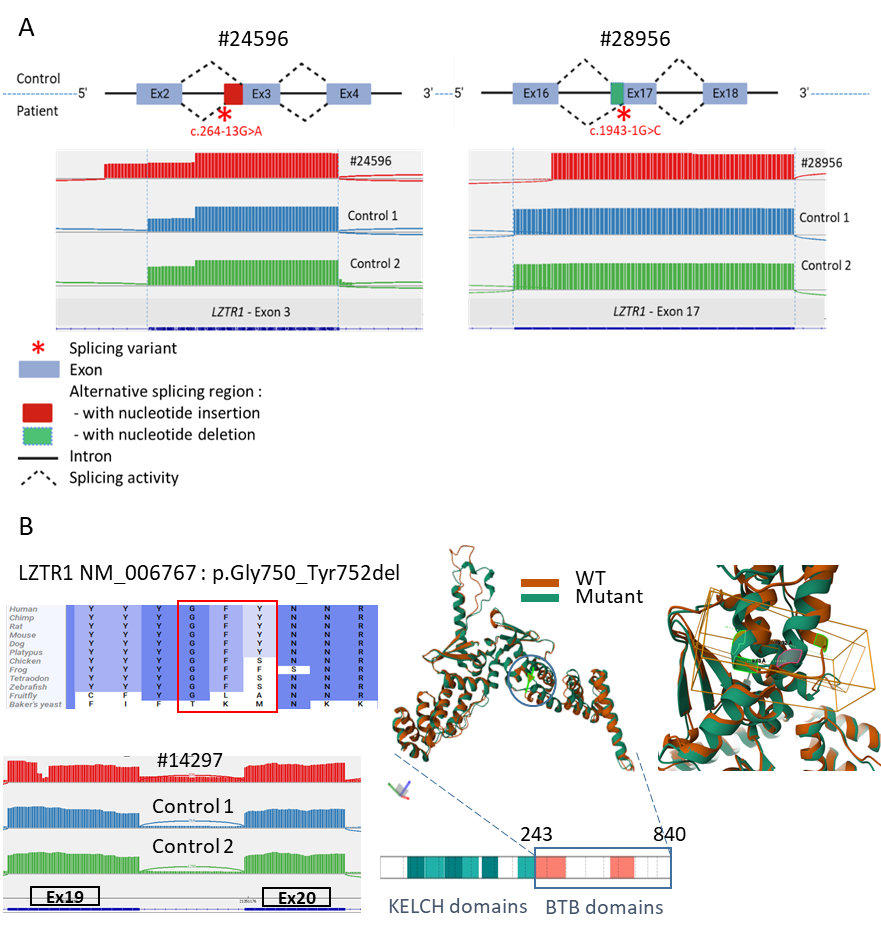

**Figure S4: SNP array profiles of 10 ALL with allelic imbalance of the *LZTR1* variant**. Somatically aquired copy-neutral loss of heterozygosity (CN-LOH) of the chromosome 22 was evidenced in 9/10 patients with *LZTR1* allelic imbalance (VAF>60%). CN-LOH was segmental (s) in 3 cases (breakpoints are indicated by red arrows), whereas it involved the whole chromosome (w) in 6 cases including 2 with duplicated hypodiploid ALL. The remaining patient had hyperdiploid ALL with chromosome 22 triploidy. NA: not available, GL: germline, S: somatic, VAF: variant allele frequency

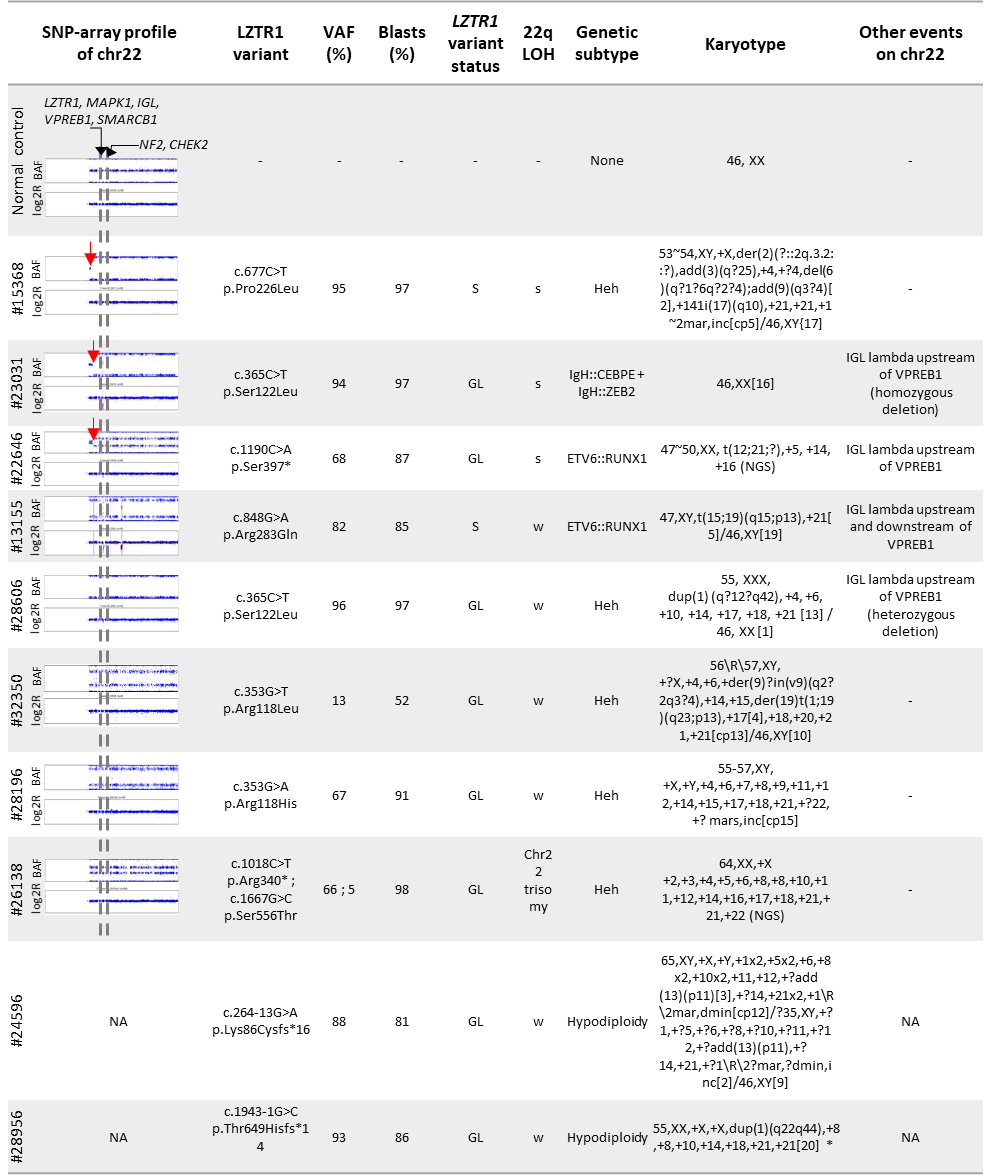

*** At relapse, cytogenetic analysis revealed a hypodiploid karyotype: 29,XX, +8, +10, +14, +18, +21[15] /46,XX[7], leading to the diagnosis of hypodiploidy.**

**Figure S5: 22q structural alterations involving the VPREB1 locus in ALL.** Overview of copy number alterations (CNA) and copy-neutral loss of heterozygosity (CN-LOH) on chromosome 22 identified by SNP array in ALL samples with 22qCN-LOH (*LZTR1*^mut^-ALL, n = 8) and in *LZTR1* wild-type controls (*LZTR1*^wt^-ALL, n = 30). Deletions, duplications, and 22qCN-LOH are shown in red, blue, and yellow, respectively. CNAs were mapped to chromosome 22 using the UCSC Genome Browser (hg19). Zoomed views highlight the VPREB1 locus within the immunoglobulin lambda variable (IGLV) gene cluster.

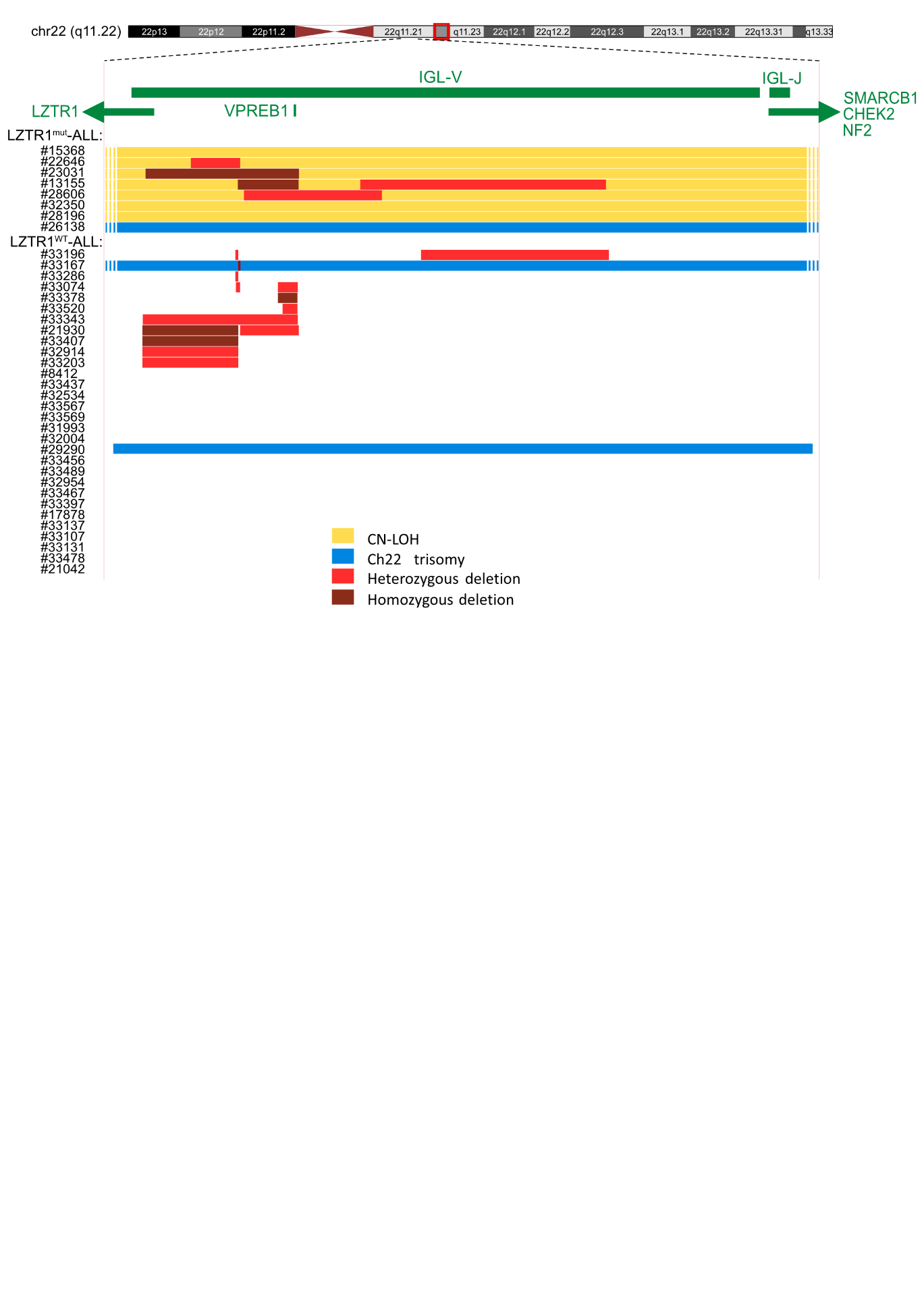

**Figure S6: RNA analysis in ALL patients. A.** Comparative analysis of variant allele frequency (VAF %) in matched DNA and RNA samples from three individual patients (#14297, #28973, #24186). For each patient, identical variants were analyzed at diagnosis (circles) and remission (triangles). Variant identifiers are indicated below each panel. Lines connect matched DNA and RNA measurements for the same variant and time point. **B.** Distribution of RNA VAF% in the WT cohort (N = 149) compared with the MT group (N = 3). Boxplots represent median and interquartile range; whiskers indicate minimum–maximum values, and crosses denote the mean. Statistical significance was assessed using a Wilcoxon rank-sum test (p = 0.003138). **C.** Differential gene expression between *LZTR1*^mut^-BCP-ALL and *LZTR1*^wt^ -BCP-ALL samples across major genetic subgroups. Volcano plots showing differential expression analysis between *LZTR1*-mutated (mut) and *LZTR1* wild-type (wt) BCP-ALL cases within each major genetic subgroup: Hyperdiploid (5 vs 10), *ETV6::RUNX1* (3 vs 6), *PAX5*-altered (1 vs 2), and *ZNF384*-rearranged (1 vs 2). Each dot represents one gene. The x-axis shows the log₂(fold-change) and the y-axis the –log₁₀(q-value). No statistically significant differences were observed.

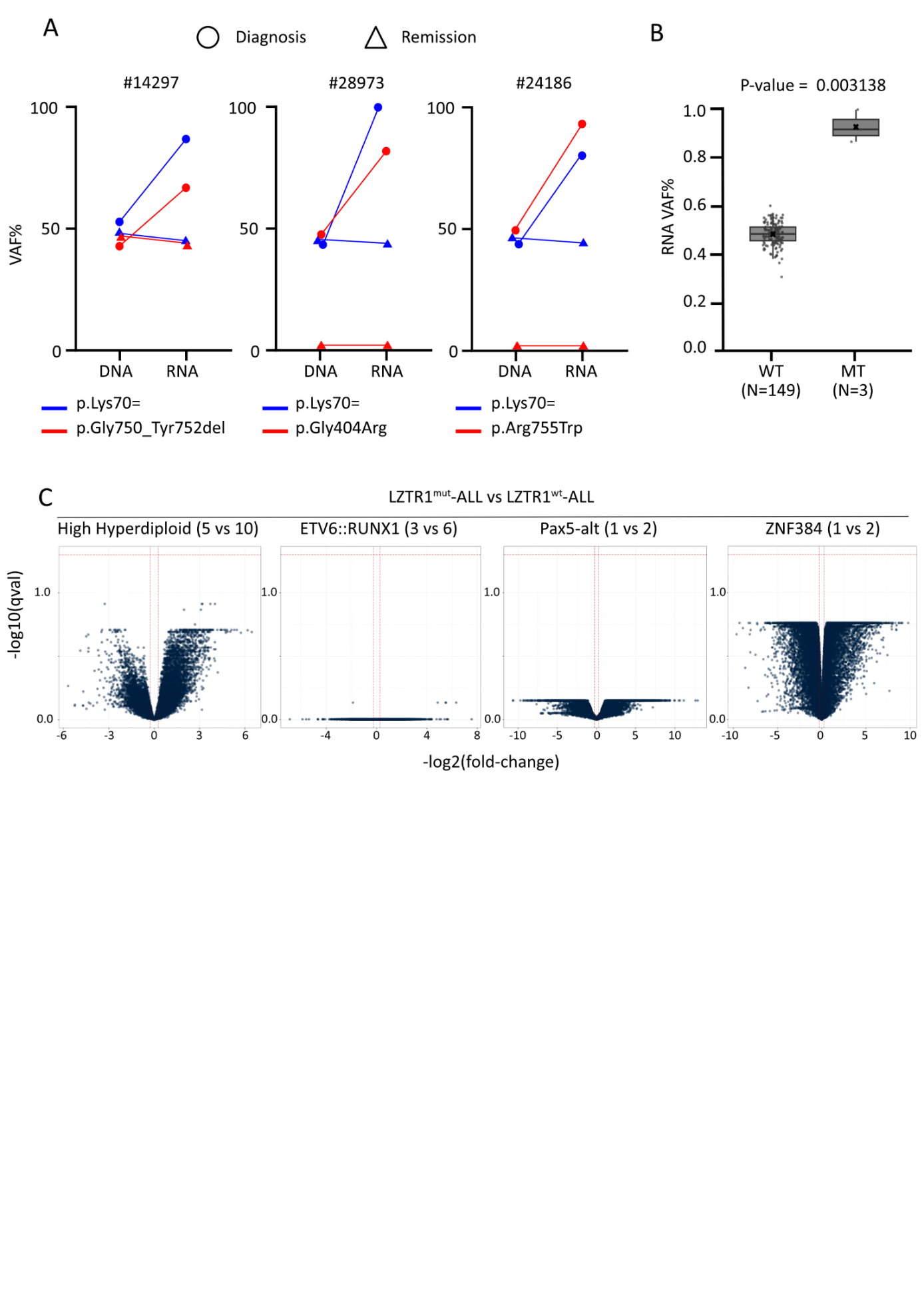

**Figure S7: Clonal architecture at diagnosis of LZTR1^mut^-ALL patients without relapse**. Fishplots represent the clonal composition of *LZTR1*-mutated ALL samples. The genetic subtype is shown in blue, RAS signaling–related clones in orange, and additional subclones in distinct colors according to their functional category. Patients carrying a germline *LZTR1* variant are highlighted with a yellow background, while those with a somatic *LZTR1* variant are shown with a grey background. Additional variants and genetic subgroups are listed next to each corresponding fishplot.

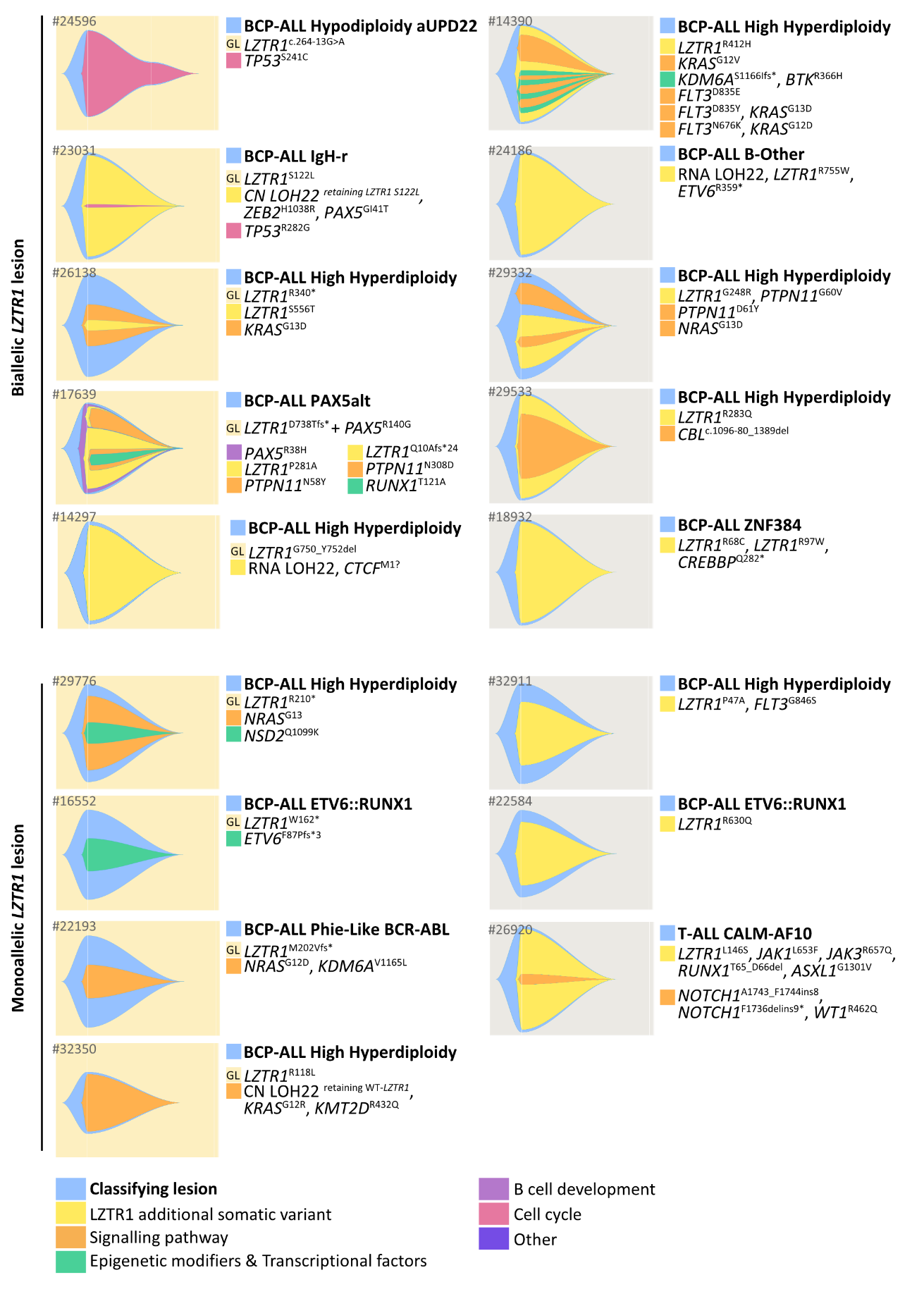

**Figure S8: Events observed during the clinical follow-up of biallelic LZTR1^mut/mut^ patients (N=19).** Each horizontal light blue bar corresponds to the follow up duration of an individual patient from diagnosis. Clinical events are indicated as follows: relapses are marked with red bars, deaths with black ǂ, and hematopoietic stem cell transplants with Δ.

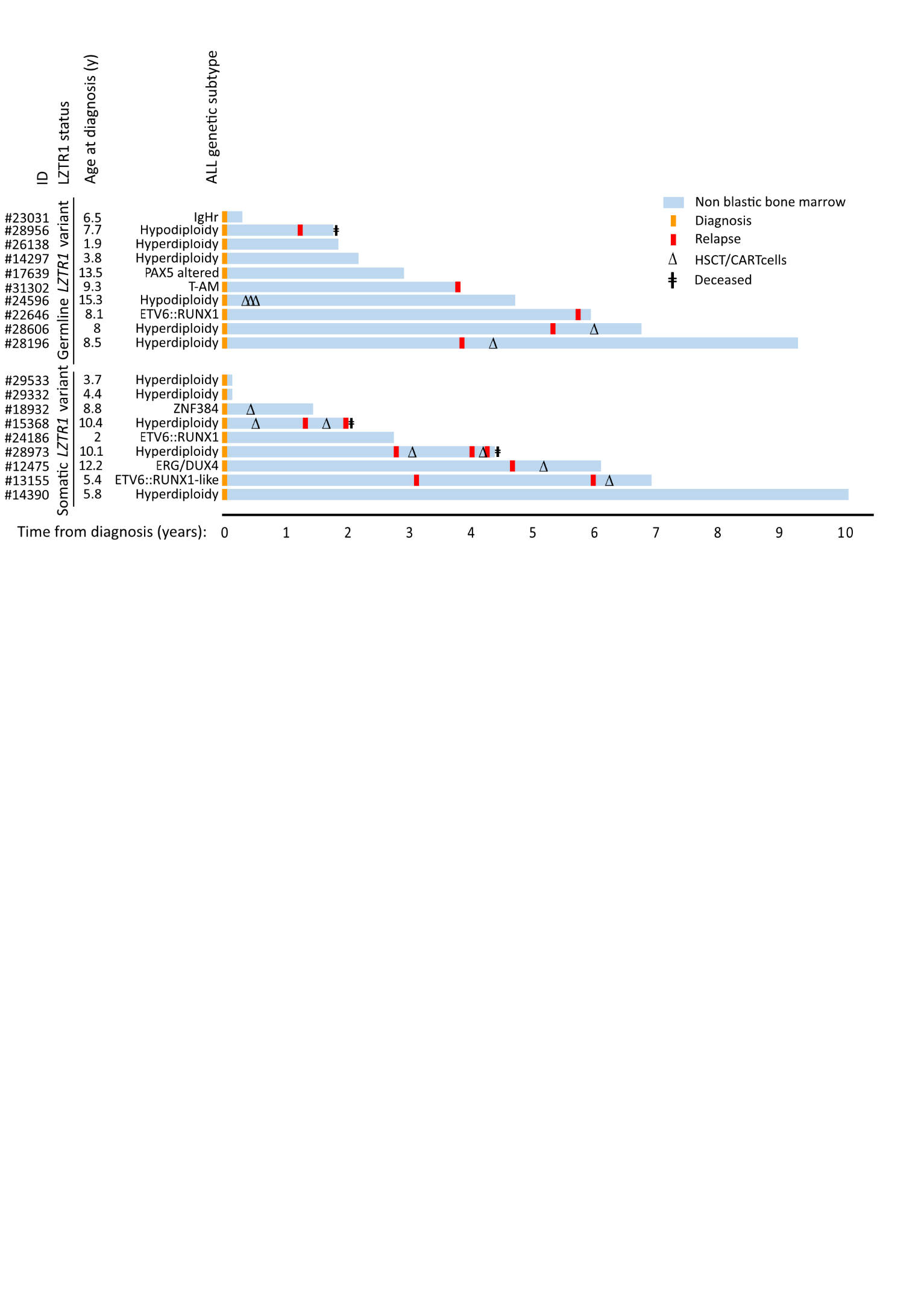

**Figure S9: Physiological transcriptomic expression of LZTR1, NF1, and LZTR1 targets genes in hematopoietic cells. A.** Bulk RNA-seq analysis of LZTR1, NF1, and RAS-pathway genes (HRAS, KRAS, NRAS, MRAS, RIT1, AXL, EGFR) across fetal and pediatric hematopoietic stem and progenitor cell (HSPC) subsets and mature lineages. Expression levels are shown as transcripts per million (TPM). **B.** Single-cell RNA-seq (scRNA-seq) analysis of the same gene set across hematopoietic populations including HSPCs, lymphoid (B and T cells), erythroid, and myeloid (granulocytes/monocytes) lineages from the DISCO single-cell human hematopoietic atlas (10x Genomics platform; available at https://disco.bii.a-star.edu.sg/atlas/bone_marrow). Log-normalized expression values are visualized on UMAP plots. Abbreviations: HSPC, hematopoietic stem and progenitor cell; CMP, common myeloid progenitor; GMP, granulocyte-macrophage progenitor; MEP, megakaryocyte-erythroid progenitor.

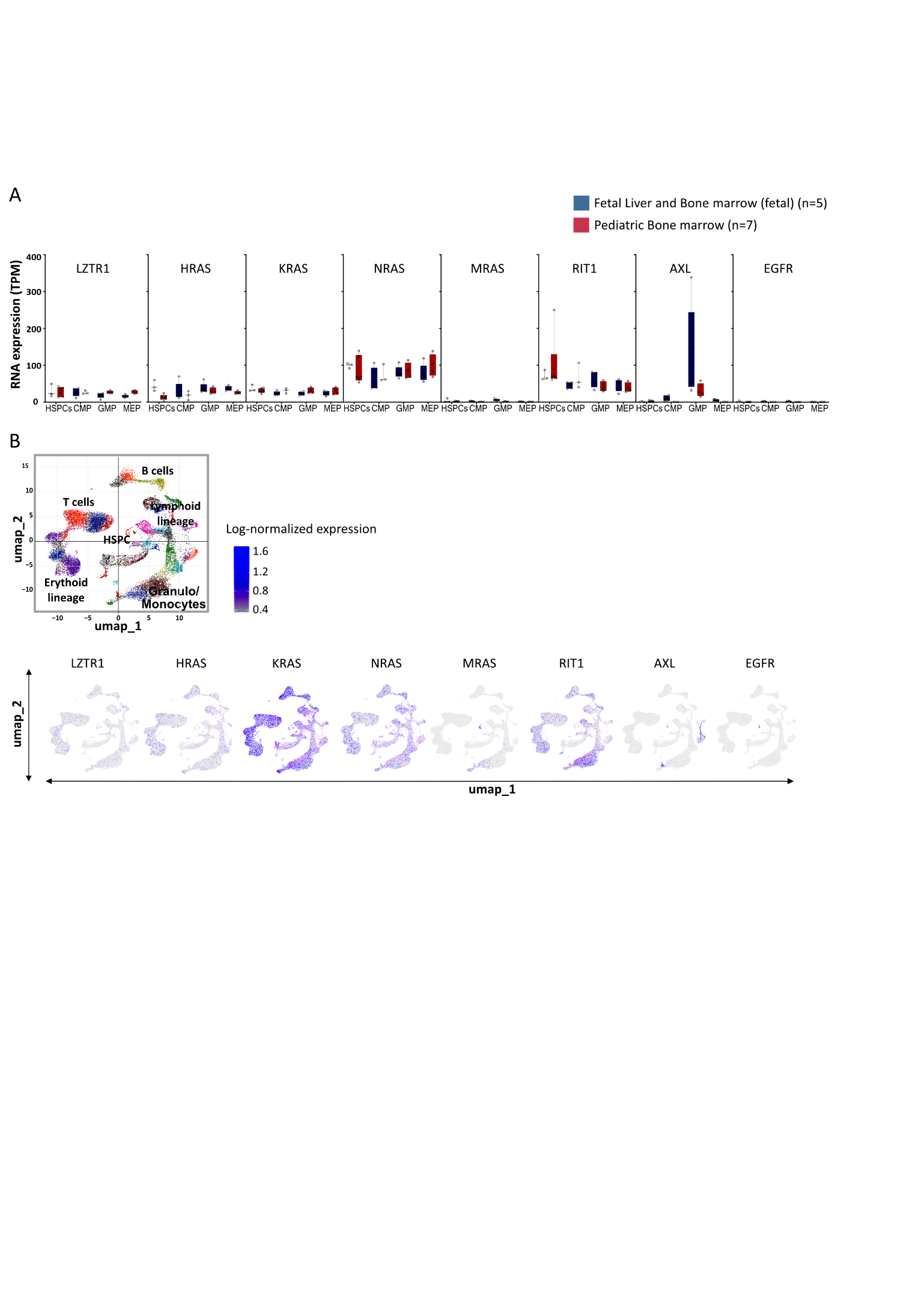

**Figure S10: Expression of LZTR1 and its putative targets in myeloid and lymphoid cell lines**. Protein expression assessed by capillary immunoassay on a Jess® apparatus (upper panel). Target protein expression (top) and total protein expression (bottom) are shown. Histograms showing quantitative expression data normalized on total protein expression using the Total Protein Detection Module® (lower panel).

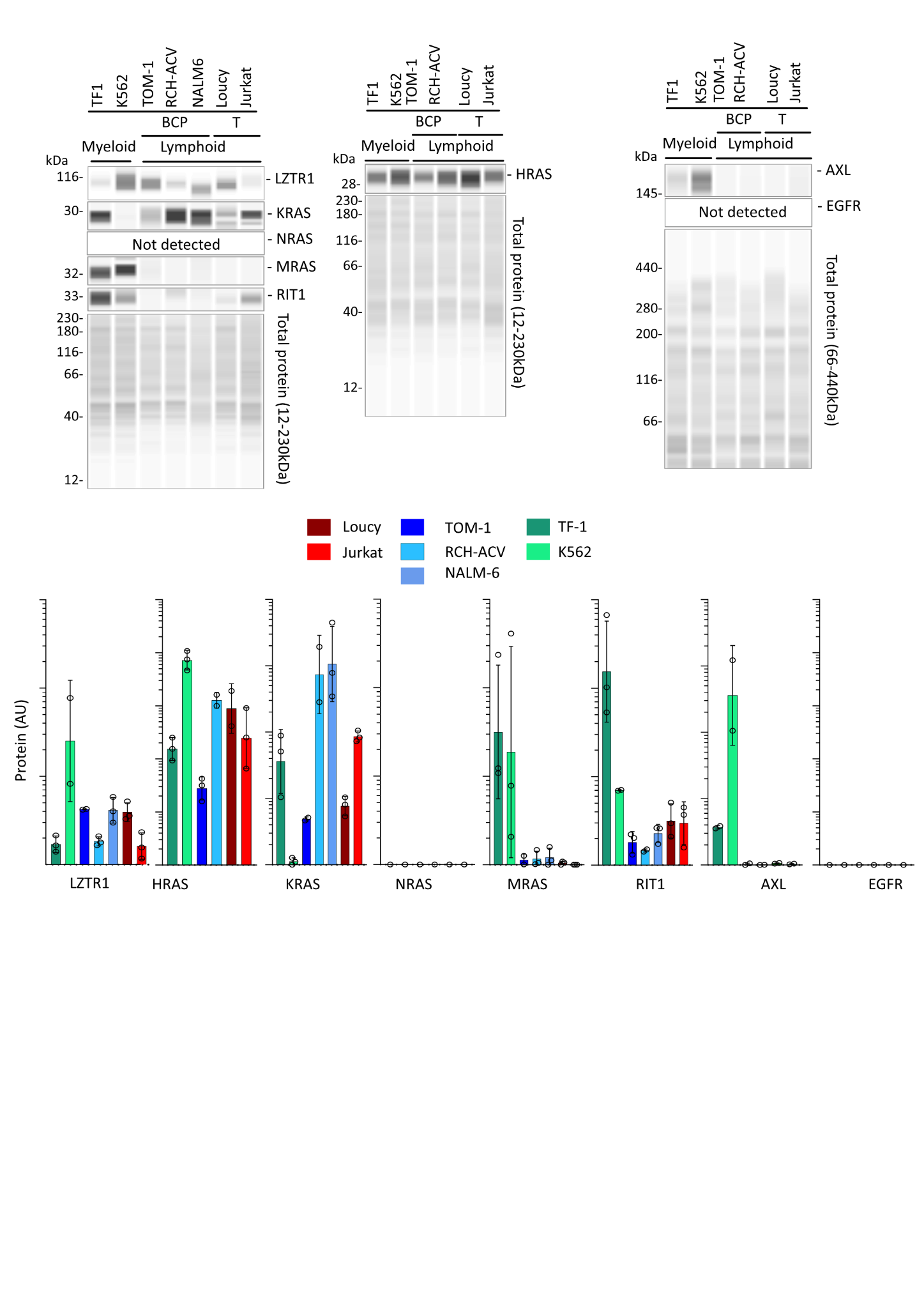

**Figure S11: Effect of LZTR1 inactivation in cell lines. A.** RAS Protein dosage in LZTR1^+/+^ and LZTR1^-/-^ cell lines. **B.** MAPK and AKT signalization in LZTR1^+/+^ and LZTR1^-/-^ cell lines at baseline and after stimulation (TF-1: GM-CSF; Jurkat: FBS stimulation). **C.** Effect of LZTR1 alteration on RAS in BCP-ALL samples, stratified by LZTR1 status (+/+, +/-, -/-); GM-CSF, granulocyte–macrophage colony-stimulating factor; FBS: fetal bovine serum

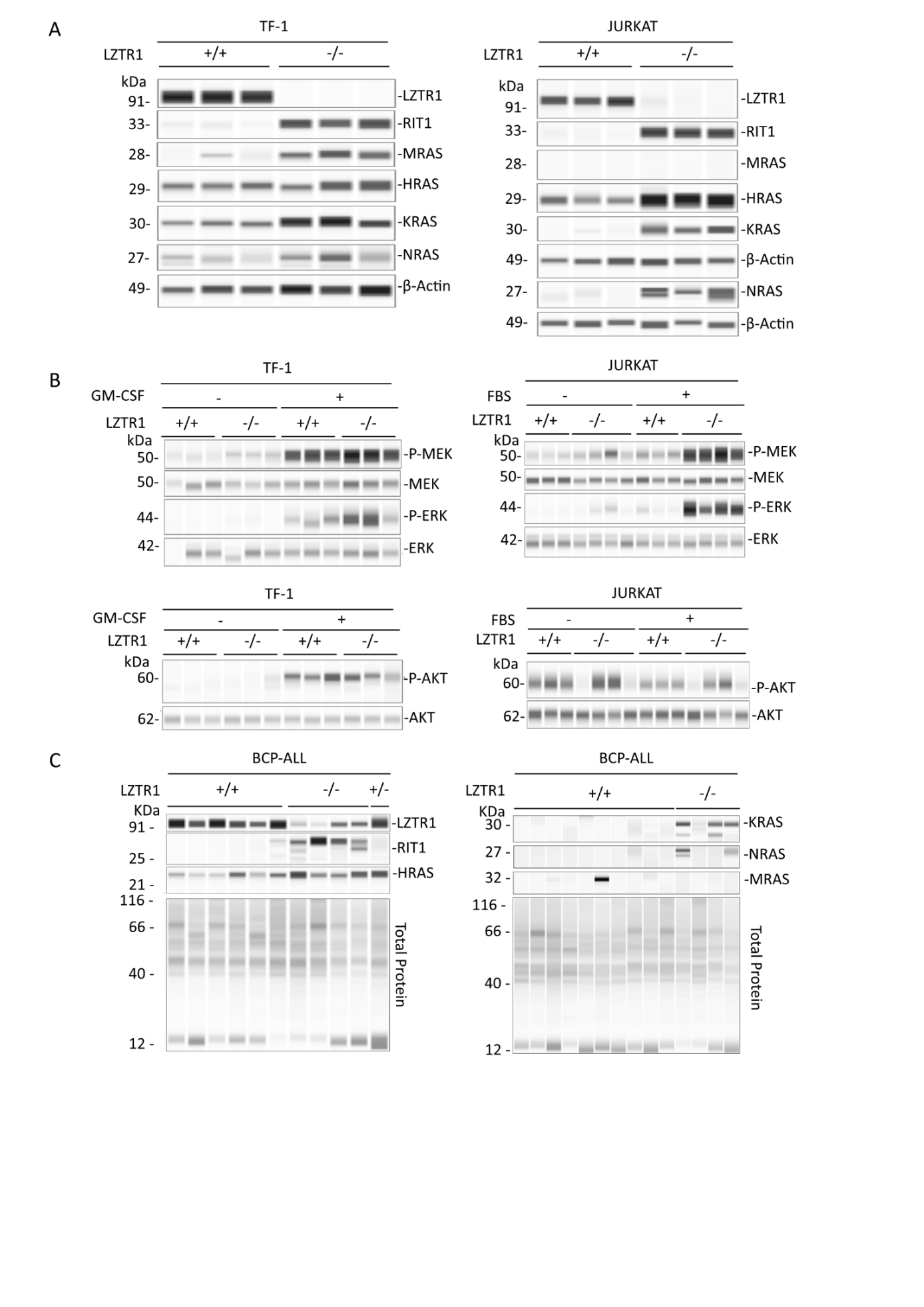

**Figure S12: Effect of LZTR1 knock-out on RAS protein expression in lymphoid cell line.** Protein levels of HRAS, KRAS, NRAS, and RIT1 were quantified and normalized to actin. Data are shown for wild-type (LZTR1+/+), heterozygous (+/−), and knock-out (−/−) clones. Dots indicate individual biological replicates. Statistical significance was assessed by one-way ANOVA with post hoc multiple comparisons test. ns, not significant; **p < 0.01; ***p < 0.001.

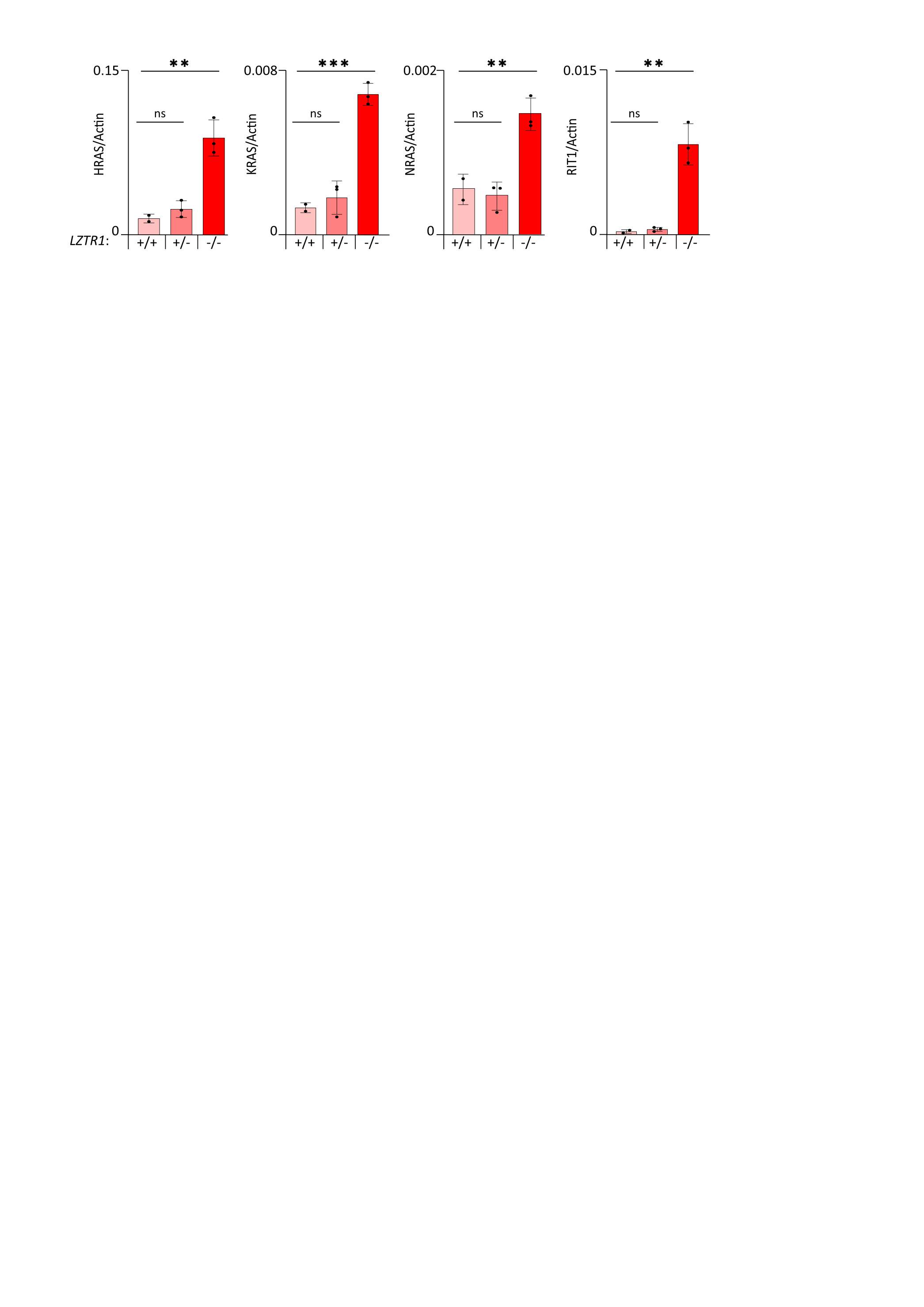

**Figure S13: Transcript expression of LZTR1, its targets and receptor tyrosine kinases in genetic subtypes of BCP-ALL. A**. Bulk total RNA-seq expression (transcripts per million, TPM) of LZTR1 across major genetic BCP-ALL subgroups (n = 204 LZTR1^WT^ cases) **B.** Bulk RNA-seq expression (TPM) of **KRAS, NRAS, MRAS, HRAS**, and **RIT1** across all LZTR1^WT^ BCP-ALL samples (n = 204). **C.** Expression levels (TPM) of the same RAS-family members across major genetic subgroups of LZTR1^WT^ BCP-ALL (n = 204). **D.** Expression levels (TPM) of **EGFR** and **AXL** transcripts across LZTR1^WT^ BCP-ALL genetic subgroups. **E.** Comparison of EGFR and AXL transcript levels between LZTR1^WT^ and LZTR1^mut^ BCP-ALL samples. ns, not significant.

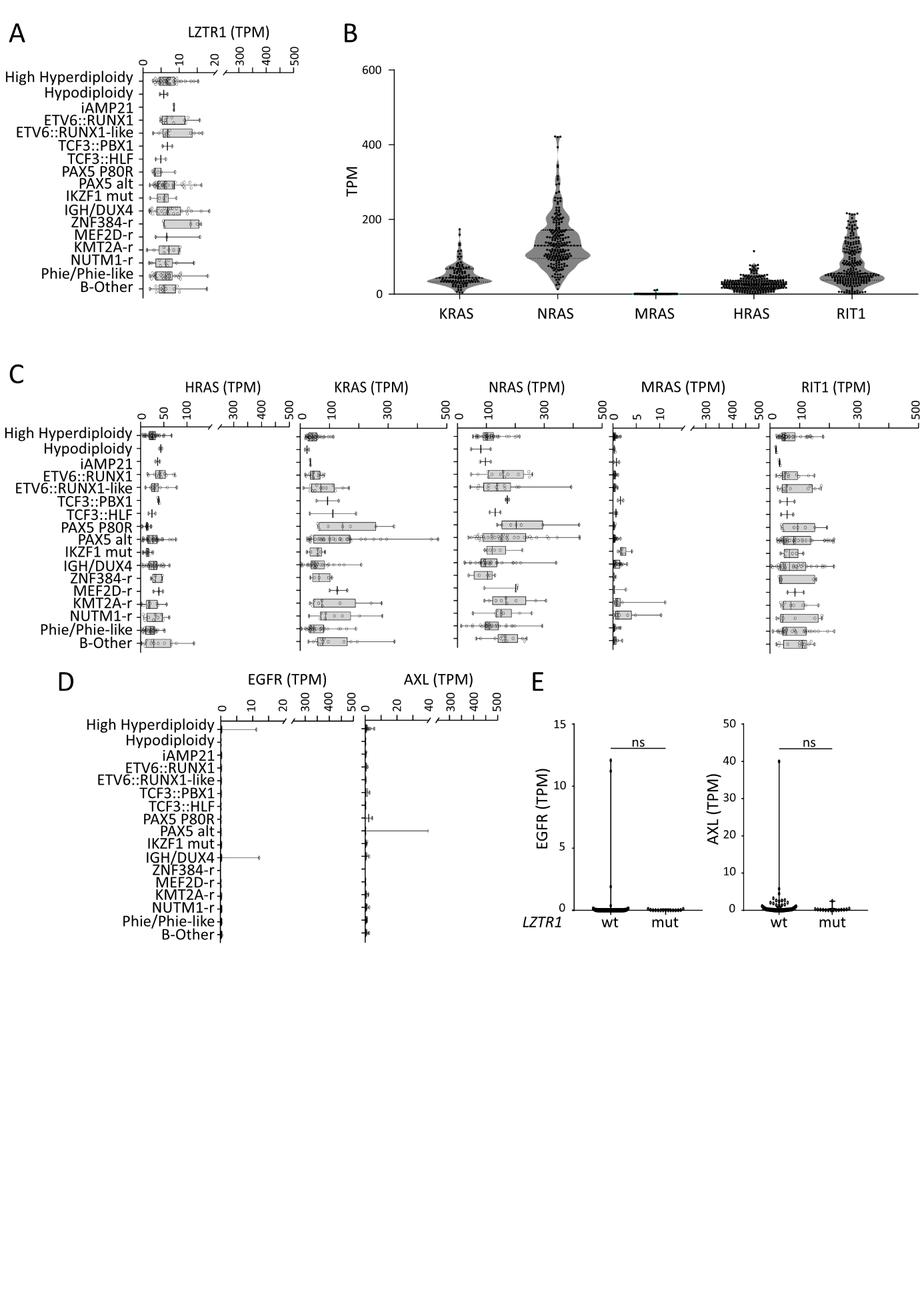
