## Supplemental Methods for "LZTR1 functions as a two-hit tumor suppressor in childhood acute lymphoblastic leukemia"

**Supplementary Material and Methods**

**Population frequency study**

General population data were obtained from the Genome Aggregation Database (gnomAD v2.1.1, non-cancer subset; RRID:SCR_014964), downloaded on January 13, 2023, using the Ensembl canonical *LZTR1* transcript (ENST00000215739.8; RRID:SCR_002344). Variants selection was adapted from Deng et al. 2022(1) with adaptations. Briefly, loss-of-function (LoF) variants — including nonsense, frameshift, and canonical splice-site (±1 or ±2) changes — were extracted in a first dataset, and missense variants and in-frame deletions in a second dataset. Variants flagged for dubious quality or with an allele frequency above 5×10⁻⁴ in any subpopulation were excluded.

For each dataset, the average number of individuals genotyped in the gnomAD non-cancer cohort was estimated by summing the “allele number” values, dividing the total by the number of variants, and halving to account for diploidy. The sum of the “allele count” values represented the total number of LZTR1 variants in the general population.

**Curated *LZTR1* variant dataset from literature: Noonan syndrome and schwannomatosis**

Variants in *LZTR1* associated with Noonan syndrome were compiled through a literature review of studies published between 2014 and 2025 (see Supplementary References(2–29)). Publications were identified using PubMed (RRID:SCR_004846) and screened for reported variants. Extracted variants were curated to remove duplicates and standardized according to Human Genome Variation Society (HGVS) nomenclature where applicable. *LZTR1* variants associated with schwannomatosis were obtained from Uliana et al.2024(30).

**DNA sequencing**

DNA libraries were prepared using SureSelect kits (XTHS2 or QXT; Agilent Technologies, Santa Clara, CA, USA), and target enrichment was performed with either a custom gene panel or the SureSelect Human All Exon V7 kit for whole exome sequencing. Sequencing was carried out using Illumina paired-end technology (2 × 150 bp; Illumina, San Diego, CA, USA) on a NextSeq 500 or NextSeq 2000 with the High Output Kit v2. A minimum coverage depth of 20× was achieved for all target regions.

Bioinformatic analysis was performed using the BWA Enrichment v2.1 pipeline (BaseSpace®, Illumina, RRID:SCR_010910). Variant calling was conducted with VarScan v2.3.5 (RRID:SCR_006849), and variant interpretation was carried out using Alissa Interpret v5 (Agilent). All reported variants were manually reviewed in Alamut® Visual.

The custom gene panel included the whole coding sequence and flanking regions of the following genes: *LZTR1, RIT1, MRAS*, and 82 genes recurrently altered in acute lymphoblastic leukemia (ALL): *ABL1, ABL2, ARID4B, BLNK, BRAF, BTG1, BTLA, CBL, CCND3, CD19, CDKN1B, CDKN2A, CDKN2B, CEBPA, CEBPB, CEBPD, CEBPG, CREBBP, CRLF2, CSF1R, EBF1, EP300, EPOR, ERG, ETV6, FBXW7, FLT3, FPGS, GATA3, IDH1, IDH2, IKZF1, IKZF2, IKZF3, IL2RB, IL7R, JAK1, JAK2, JAK3, KDM6A, KIT, KMT2A, KMT2D, KRAS, MLH1, MSH2, MSH6, MYB, MYC, NF1, NOTCH1, NPM1, NR3C1, NR3C2, NSD2, NRAS, NT5C2, NTRK3, PAG1, PAX5, PBX1, PDGFRA, PDGFRB, PMS2, PRPS1, PRPS2, PTEN, PTK2B, PTPN11, RB1, SETD2, SH2B3, STAT5A, STAT5B, SYK, TBL1XR1, TP53, TYK2, USP9X, VPREB1, WT1,* and *ZEB2*.

**Whole RNA sequencing**

Libraries were prepared with NEBNext Ultra II Directional RNA Library Prep Kit, according supplier recommendations. Paired-end 100-bp reads sequencing was performed on a NovaSeq platform. Image analysis and base calling was performed using Illumina Real Time Analysis (3.4.4) with default parameters.

*Quantification of gene expression*: STAR (RRID:SCR_004463) was used to obtain the number of reads associated to each gene in the Gencode v31 annotation (restricted to protein-coding genes, antisense and lincRNAs). Raw counts for each sample were imported into R statistical software. Extracted count matrix was normalized for library size and coding length of genes to compute TPM expression levels.

*Unsupervised analysis:* The Bioconductor edgeR package (RRID:SCR_012802) was used to import raw counts into R statistical software, and compute normalized log2 CPM (counts per millions of mapped reads) using the TMM (weighted trimmed mean of M-values) as normalization procedure. The normalized expression matrix from a custom gene list was used to classify the samples according to their gene expression patterns using principal component analysis (PCA), hierarchical clustering and consensus clustering. PCA was performed by FactoMineR::PCA function with “ncp = 10, scale.unit = FALSE” parameters. Hierarchical clustering was performed by stats::hclust function (with euclidean distance and ward.D method). Consensus clustering was performed by ConsensusClusterPlus::ConsensusClusterPlus function to examine the stability of the clusters. We established consensus partitions of the data set in K clusters (for K = 2, 3, . . . , 8), on the basis of 1,000 resampling iterations (80% of genes, 80% of sample) of hierarchical clustering, with euclidean distance and ward.D method. Then, the cumulative distribution functions (CDFs) of the consensus matrices were used to determine the optimal number of clusters (K = 3 for instance), considering both the shape of the functions and the area under the CDF curves.

*Differential expression analysis:* The Bioconductor edgeR package was used to import raw counts into R statistical software. Differential expression analysis was performed using the Bioconductor limma package (RRID:SCR_010943) and the voom transformation. To improve the statistical power of the analysis, only genes expressed in at least one sample (TPM≥0.3) were considered. A qval threshold of ≤0.05 and a minimum fold change of 1.2 were used to define differentially expressed genes.

**Targeted RNA sequencing**

RNA libraries were prepared using the SureSelect XTHS2 RNA kit (Agilent Technologies) and sequenced on a NextSeq 500® system using the High Output Kit v2 with 2 × 150 bp paired-end reads (Illumina). Bioinformatic processing used the RNA_SEQ_UMI 1.1.1 workflow on the MOABI-APHP (Multi Omics Analysis & BIoInformatic-APHP) platform. Gene quantification was performed with Salmon v0.7.2 (RRID:SCR_017036), and splice junction analyses with STAR aligner.

*Assessment of LZTR1 loss of heterozygosity at the RNA level: LZTR1* was sequenced by NGS in DNA and RNA extracted from the same leukemia and the variant allele frequency (VAF) of the *LZTR1* variant was calculated for RNA and DNA. An imbalance greater than 25% in RNA compared with the corresponding DNA VAF was considered significant and classified as "RNA LOH22". Allelic imbalance was also assessed in a group of informative LZTR1^wt^ ALL (n=149), using heterozygous benign variant within the *LZTR1* coding sequence (c.210G>A p.K70= and c.1683C>T p.R561=).

**Copy number alteration (CNA) screening**

CNA were screened using either SNP array or NGS genotyping.

SNP array analysis were performed on genomic DNA using the Infinium OmniExpress-700K SNP array (Illumina). Intensity data were processed with GenomeStudio (Illumina) to obtain log₂ ratio and B-allele frequency (BAF) values. Copy-number alterations were inferred from log₂ ratio deviations, and copy-neutral loss of heterozygosity (CN-LOH) was defined as regions showing a near-zero log₂ ratio (diploid copy number) associated with a shift in BAF from the expected heterozygous value of 0.5. CN-LOH regions were visually confirmed by concurrent examination of log₂ ratio and BAF plots, including mosaic patterns when present.

NGS-based genotyping was performed by analyzing single-nucleotide polymorphisms (SNPs) located in genes distributed along chromosome 22 (*LZTR1, VPREB1, SMARCB1, IL2RB, EP300, RAC2,* and *SH3BP1*). For each patient, heterozygous and homozygous SNPs were extracted and analyzed. CN-LOH was suspected when ≥85% of informative SNPs were homozygous across at least three distinct genes. When fewer than 20 informative SNPs were available, CN-LOH assessment was considered inconclusive due to limited data. In both situations—either when CN-LOH criteria were fulfilled or when too few informative SNPs were detected—results were systematically verified using SNP array to confirm or exclude copy-number alterations or loss of heterozygosity.

**Co-mutation enrichment analysis (RAS pathway)**

Co-mutation analyses were performed on genes involved in the RAS signaling pathway, including *KRAS, NRAS, PTPN11, NF1, FLT3, CBL*, and *LZTR1*.

For each gene and each patient (whole cohort, N=1587), VAFs from multiple variants affecting the same gene were aggregated (cumulative VAF). Mutations were then binarized using a predefined threshold, considering a gene as mutated when the cumulative VAF exceeded 20% (VAF > 20%). This resulted in a binary matrix representing the mutation status of each gene across patients.

Observed co-mutation counts were computed using matrix multiplication. Diagonal elements correspond to the number of patients harboring mutations in each gene, while off-diagonal elements represent the number of patients with co-occurring mutations for each pair of genes.

Expected co-mutation counts under the assumption of independence were estimated based on marginal mutation frequencies.

To quantify enrichment or depletion of co-mutations, the observed-to-expected ratio was transformed as:

$$\log2\left( \frac{O+0.5}{E+0.5} \right)$$

A pseudo-count of 0.5 was added to ensure numerical stability and avoid division by zero for rare events. Negative values indicate fewer co-mutations than expected (consistent with mutual exclusivity), whereas positive values indicate enrichment (co-occurrence).

Only the lower triangular portion of the symmetric matrix was retained for visualization to avoid redundancy. Heatmaps were generated using a diverging color scale centered at zero.

**Cell culture conditions**

Cell lines (Table S1) were maintained in RPMI 1640 medium supplemented with 10% fetal bovine serum (FBS; Gibco), 1% penicillin/streptomycin, and 5% CO₂ at 37 °C. TF-1 cells additionally received 1% sodium pyruvate and 5 ng/mL recombinant human GM-CSF (PeproTech). All cell lines were routinely tested for Mycoplasma contamination. Cell line identity was checked before and after CRISPR/Cas9 KO by comparing the mutational profiles obtained from targeted sequencing with publicly available genomic data from the Cancer Dependency Map (DepMap, Broad Institute). Consistency with the expected genotypes was confirmed for each cell line (<https://depmap.org/portal/>).

**Signaling assays**

Cells were maintained in their respective growth media until 24 hours before stimulation, then subjected to serum deprivation (RPMI 1640 containing 1% fetal bovine serum) to reduce basal signaling activity. TF-1 cells were stimulated with 5 ng/mL recombinant human GM-CSF (PeproTech) for 15 minutes at 37 °C. Jurkat cells were stimulated for 10 minutes either with a T-cell activation cocktail (50 ng/mL PMA and 1 µM ionomycin) or by re-addition of 10% FBS to serum-deprived cultures. Cells were immediately harvested on ice, washed with PBS, and processed for downstream protein analyses

**Generation of CRISPR/Cas9 knockout cell lines**

*CRISPR/Cas9 editing:* Gene inactivation was achieved using the ‘all-in-one’ Cas9–GFP plasmid (CMV-CG-Cas9-2A-tGFP, Sigma-Aldrich). sgRNAs targeting LZTR1 were designed using the Synthego CRISPR Design Tool, Broad Institute GPP Designer, and Invitrogen TrueDesign Genome Editor. A non-targeting control gRNA targeting the AAVS1 locus (*PPP1R12C* gene) was used as control.

*Plasmid preparation:* Plasmids were amplified in E. coli One Shot® TOP10 cells and purified using the NucleoBond™ Xtra Midi EF kit (Macherey-Nagel). Bacteria were grown in Kanamycin-containing medium (Invitrogen™ imMedia™ Kan Liquid).

*Nucleofection and clone isolation:* Nucleofections were performed with 2 µg of plasmid using a Lonza Nucleofector 2b or 4D system and the Amaxa Human SE/SF Cell Line kits, following manufacturer programs (CL-120, T-001). Cells were recovered for 48 h and single-cell clones isolated using WOLF or Aria II FACS based on GFP and viability dyes (propidium iodide or Zombie Violet). Single cells were sorted into 96-well plates containing complete RPMI and cultured for two weeks.

*Clone validation:* Genomic DNA was extracted and CRISPR edits confirmed by PCR and Sanger sequencing. LZTR1 inactivation was verified by capillary immunoassay (Jess®, ProteinSimple). Whole-exome and targeted DNAseq of RAS pathway genes were performed to evaluate the possible off-target effect and document the presence of RAS-MAPK pathway mutations that could potentially interfere with the results (Supp Table 6).

**Protein preparation and immunoassays**

*Protein extraction*: cells were lysed using M-PER (Mammalian Protein Extraction Reagent), supplemented with 1 mM PMSF (Thermofisher), one tablet of PhosSTOP™ (Sigma-Aldrich) per 10 mL and protease cocktail inhibitor (P8340, Thermo Fisher Scientific), incubated for 10 min on ice with agitation. Lysates were cleared by centrifugation (13000 rpm, 15min, 4°C). The proteins were quantified and normalized with Bioanalyzer 2100 Kit protein230.

*Capillary immunoassay*: Immunodetection was performed using the Jess® automated capillary electrophoresis system (ProteinSimple), using the 12–230 kDa or the 66-440 kDa capillary cartridge according to manufacturer’s protocol. Antibody and reagent details are in Table S1. The optimal antibody dilution was assessed by antibody titration prior to use on cell-lines. Normalization was performed on β -actin or on the total protein amount of the sample, using Total Protein Detection Module®.

At least two replicates were performed for each condition, unless otherwise stated. Each experiment was repeated at least twice to ensure technical reproducibility.

**Pull-down assay**

RAS-GTP pull-down was performed using the GST–Raf1-RBD kit (Ras Pull-Down and Detection Kit, Thermo Fisher Scientific, #16117) following manufacturer’s protocol. Briefly, 1×10⁷ cells were processed per condition. After washing, bound proteins were eluted and denatured at 95°C for 5 min. Total RAS was quantified by capillary immunoassay in the eluate and in the input lysates. Total RAS in input lysates was used for normalization.

Each condition was tested in duplicate

**Statistical analysis and graphical representation.**

Statistical analyses were performed using GraphPad Prism version 10.0.0 for Windows, GraphPad Software, Boston, Massachusetts USA, [www.graphpad.com](http://www.graphpad.com). Two-tailed unpaired Student’s t-tests were used for comparisons between two groups. For multiple group comparisons, one-way or two-way ANOVA was applied as appropriate, followed by post hoc multiple comparison tests. **** equals p value ≤ 0.0001. p < 0.05 was considered statistically significant.

Fishplots were created with the chrisamiller/fishplot (v.0.5 ; RRID:SCR_016839) package using RStudio (2022.02.0+443; RRID:SCR_000432).

Lollipop plots were generated using ProteinPaint (https://proteinpaint.stjude.org), a visualization tool developed by St. Jude Children’s Research Hospital (Zhou et al., Nat Genet, 2016).

Event-free survival, overall survival, and cumulative incidence of relapse were estimated in both the overall cohort and the subgroup of low-risk leukemia patients over a 12-year follow-up. Kaplan–Meier curves and cumulative incidence plots were generated with 95% confidence intervals (Cis) and p-values. Confidence intervals were computed using Greenwood’s formula with the log(–log) transformation.

Statistical analyses were conducted using R with the survival, survminer, dplyr, and coxphf packages. Differences in survival between groups were assessed using the log-rank test. Effect sizes were estimated as hazard ratios (HRs) with 95% confidence intervals derived from Cox proportional hazards models.

All statistical tests were two-sided, with a significance threshold of p < 0.05.

**Summary of analyses per sample.**

A complete overview of the analyses performed for each patient and control sample is provided in Table S7.

**Supplementary references**

1.	Deng F, Evans DG, Smith MJ. Comparison of the frequency of loss-of-function LZTR1 variants between schwannomatosis patients and the general population. Hum Mutat. 2022 Jul;43(7):919–27. doi:10.1002/humu.24376 PubMed PMID: 35391499; PubMed Central PMCID: PMC9324957.

2.	Chen PC, Yin J, Yu HW, Yuan T, Fernandez M, Yung CK, et al. Next-generation sequencing identifies rare variants associated with Noonan syndrome. Proceedings of the National Academy of Sciences. 2014 Aug 5;111(31):11473–8. doi:10.1073/pnas.1324128111

3.	Yamamoto GL, Aguena M, Gos M, Hung C, Pilch J, Fahiminiya S, et al. Rare variants in SOS2 and LZTR1 are associated with Noonan syndrome. J Med Genet. 2015 Jun;52(6):413–21. doi:10.1136/jmedgenet-2015-103018

4.	Johnston JJ, van der Smagt JJ, Rosenfeld JA, Pagnamenta AT, Alswaid A, Baker EH, et al. Autosomal recessive Noonan syndrome associated with biallelic LZTR1 variants. Genet Med. 2018;20(10):1175–85. doi:10.1038/gim.2017.249 PubMed PMID: 29469822; PubMed Central PMCID: PMC6105555.

5.	Shamseldin HE, Kurdi W, Almusafri F, Alnemer M, Alkaff A, Babay Z, et al. Molecular autopsy in maternal–fetal medicine. Genetics in Medicine. 2018 Apr;20(4):420–7. doi:10.1038/gim.2017.111

6.	Güemes M, Martín-Rivada Á, Ortiz-Cabrera NV, Martos-Moreno GÁ, Pozo-Román J, Argente J. LZTR1: Genotype Expansion in Noonan Syndrome. Horm Res Paediatr. 2019;92(4):269–75. doi:10.1159/000502741

7.	Li X, Yao R, Tan X, Li N, Ding Y, Li J, et al. Molecular and phenotypic spectrum of Noonan syndrome in Chinese patients. Clin Genet. 2019 Oct;96(4):290–9. doi:10.1111/cge.13588

8.	Motta M, Fidan M, Bellacchio E, Pantaleoni F, Schneider-Heieck K, Coppola S, et al. Dominant Noonan syndrome-causing LZTR1 mutations specifically affect the Kelch domain substrate-recognition surface and enhance RAS-MAPK signaling. Human Molecular Genetics. 2019 Mar 15;1007–22.

9.	Nakaguma M, Jorge AAL, Arnhold IJP. Noonan syndrome associated with growth hormone deficiency with biallelic LZTR1 variants. Genet Med. 2019 Jan;21(1):260–260. doi:10.1038/s41436-018-0041-5

10.	Pagnamenta AT, Kaisaki PJ, Bennett F, Burkitt‐Wright E, Martin HC, Ferla MP, et al. Delineation of dominant and recessive forms of LZTR1-associated Noonan syndrome. Clin Genet. 2019 Jun;95(6):693–703. doi:10.1111/cge.13533

11.	Perin F, Trujillo-Quintero JP, Jimenez-Jaimez J, Rodríguez-Vázquez del Rey M del M, Monserrat L, Tercedor L. Two Novel Cases of Autosomal Recessive Noonan Syndrome Associated With LZTR1 Variants. Revista Española de Cardiología (English Edition). 2019 Nov;72(11):978–80. doi:10.1016/j.rec.2019.05.002

12.	Umeki I, Niihori T, Abe T, Kanno S ichiro, Okamoto N, Mizuno S, et al. Delineation of LZTR1 mutation-positive patients with Noonan syndrome and identification of LZTR1 binding to RAF1–PPP1CB complexes. Hum Genet. 2019 Jan;138(1):21–35. doi:10.1007/s00439-018-1951-7

13.	Bertola DR, Castro MAA, Yamamoto GL, Honjo RS, Ceroni JR, Buscarilli MM, et al. Phenotype–genotype analysis of 242 individuals with RASopathies: 18‐year experience of a tertiary center in Brazil. Am J Med Genet. 2020 Dec;184(4):896–911. doi:10.1002/ajmg.c.31851

14.	Chinton J, Huckstadt V, Mucciolo M, Lepri F, Novelli A, Gravina LP, et al. Providing more evidence on LZTR1 variants in Noonan syndrome patients. Am J Med Genet. 2020 Feb;182(2):409–14. doi:10.1002/ajmg.a.61445

15.	Ferrari L, Mangano E, Bonati MT, Monterosso I, Capitanio D, Chiappori F, et al. Digenic inheritance of subclinical variants in Noonan Syndrome patients: an alternative pathogenic model? Eur J Hum Genet. 2020 Oct;28(10):1432–45. doi:10.1038/s41431-020-0658-0

16.	Jacquinet A, Bonnard A, Capri Y, Martin D, Sadzot B, Bianchi E, et al. Oligo-astrocytoma in LZTR1-related Noonan syndrome. European Journal of Medical Genetics. 2020 Jan;63(1):103617. doi:10.1016/j.ejmg.2019.01.007

17.	Hanses U, Kleinsorge M, Roos L, Yigit G, Li Y, Barbarics B, et al. Intronic CRISPR Repair in a Preclinical Model of Noonan Syndrome–Associated Cardiomyopathy. Circulation. 2020 Sep 15;142(11):1059–76. doi:10.1161/CIRCULATIONAHA.119.044794

18.	Zhao X, Li Z, Wang L, Lan Z, Lin F, Zhang W, et al. A Chinese family with Noonan syndrome caused by a heterozygous variant in LZTR1: a case report and literature review. BMC Endocr Disord. 2021 Dec;21(1):2. doi:10.1186/s12902-020-00666-6

19.	Zhou X, Zhou J, Wei X, Yao R, Yang Y, Deng L, et al. Value of Exome Sequencing in Diagnosis and Management of Recurrent Non-immune Hydrops Fetalis: A Retrospective Analysis. Front Genet. 2021 Apr 9;12:616392. doi:10.3389/fgene.2021.616392

20.	Uludağ Alkaya D, Lissewski C, Yeşil G, Zenker M, Tüysüz B. Expanding the clinical phenotype of RASopathies in 38 Turkish patients, including the rare *LZTR1* , *RAF1* , *RIT1* variants, and large deletion in *NF1* . American J of Med Genetics Pt A. 2021 Dec;185(12):3623–33. doi:10.1002/ajmg.a.62410

21.	Dempsey E, Haworth A, Ive L, Dubis R, Savage H, Serra E, et al. A report on the impact of rapid prenatal exome sequencing on the clinical management of 52 ongoing pregnancies: a retrospective review. BJOG. 2021 May;128(6):1012–9. doi:10.1111/1471-0528.16546 PubMed PMID: 32981126.

22.	Hurni Y, Marangoni M, Garofalo G, Cassart M, Tomasi L, Vandernoot I, et al. Spontaneous resolution of nonimmune hydrops fetalis in a fetus with TP63 gene mutation and LZTR1 gene variants. Clin Case Rep. 2021 Aug;9(8). doi:10.1002/ccr3.4624

23.	Chinton J, Huckstadt V, Foncuberta ME, Perez MM, Bonetto MC, Gravina LP, et al. Challenges in genetic diagnosis, co-occurrence of 22q11.2 deletion syndrome and Noonan syndrome. American Journal of Medical Genetics Part A. 2022;188(8):2505–8. doi:10.1002/ajmg.a.62862

24.	Farncombe KM, Thain E, Barnett-Tapia C, Sadeghian H, Kim RH. LZTR1 molecular genetic overlap with clinical implications for Noonan syndrome and schwannomatosis. BMC Med Genomics. 2022 Dec;15(1):1. doi:10.1186/s12920-022-01304-x

25.	Kraoua L, Jaouadi H, Allouche M, Achour A, Kaouther H, Ahmed HB, et al. Molecular autopsy and clinical family screening in a case of sudden cardiac death reveals ACTN2 mutation related to hypertrophic/dilated cardiomyopathy and a novel LZTR1 variant associated with Noonan syndrome. Molec Gen & Gen Med. 2022 Jul;10(7). doi:10.1002/mgg3.1954

26.	De Ridder W, van Engelen B, van Alfen N. Neurological features of Noonan syndrome and related RASopathies: Pain and nerve enlargement characterized by nerve ultrasound. American Journal of Medical Genetics Part A. 2022;188(6):1801–7. doi:10.1002/ajmg.a.62714

27.	He X, Ma X, Wang J, Zou Z, Huang H, Ren J, et al. Case report: Identification and clinical phenotypic analysis of novel mutation of the PPP1CB gene in NSLH2 syndrome. Front Behav Neurosci. 2022 Sep 8;16:987259. doi:10.3389/fnbeh.2022.987259

28.	Sun L, Xie Y mei, Wang S shui, Zhang Z wei. Cardiovascular Abnormalities and Gene Mutations in Children With Noonan Syndrome. Front Genet. 2022 Jun 13;13:915129. doi:10.3389/fgene.2022.915129

29.	Wang Y, Wang W, Wang X, Xin X, Yin Y, Zhao C, et al. Abernethy malformation (Type II) presenting in a 6-day-old boy with Noonan syndrome: a case report. BMC Pediatr. 2025 Jul 12;25(1):551. doi:10.1186/s12887-025-05726-1 PubMed PMID: 40646478; PubMed Central PMCID: PMC12254992.

30.	Uliana V, Ambrosini E, Taiani A, Cesarini S, Cannizzaro IR, Negrotti A, et al. Phenotypic Expansion of Autosomal Dominant LZTR1-Related Disorders with Special Emphasis on Adult-Onset Features. Genes (Basel). 2024 Jul 13;15(7):916. doi:10.3390/genes15070916 PubMed PMID: 39062695; PubMed Central PMCID: PMC11276570.
