## Supplemental Tables 1-2 for "LZTR1 functions as a two-hit tumor suppressor in childhood acute lymphoblastic leukemia"

Adeline A. Bonnard^1^, Aurélie Caye-Eude^1,2^, Chloé Arfeuille^1,2^, Séverine Drunat^1^, Anna Dehler^3^, Fabio D. Steffen^3^, Elodie Lainey^2,4^, Damien Bodet^5^, Claire Freycon^6^, Catherine Paillard^7^, Pauline Simon^8^, Arnaud Petit^9^, Cécile Pochon^10^, Jean-Hugues Dalle^11^, Nastassja Scheidegger^3^, Beat Bornhauser^3^, André Baruchel^11^, Marion Strullu^2,11^, Yoann Vial^1^, Hélène Cavé^1,21^Service de Génétique du Développement, Hôpital Robert Debré, AP-HP – Université Paris-Cité, Paris, France

^2^INSERM U1360, Institut Gustave Roussy, Villejuif, France.

^3^Oncology department, University Children's Hospital Zurich, Switzerland

^4^Service d’Hématologie Biologique, Hôpital Robert Debré, AP-HP – Université Paris-Cité, Paris, France

^5^Unité d'Onco-Hémato-Pédiatrique, CHU de Caen, Caen, France

^6^Pediatrics, CHRU La Tronche, Grenoble, France

^7^Department of Pediatric Hematology and Oncology, CHU Hautepierre, Strasbourg, France.

^8^Department of Pediatric Hematology-Oncology CHRU Besançon, Besançon, France.

^9^Hôpital Armand-Trousseau, APHP, Université Paris-Sorbonne, Paris, France

^10^Department of Pediatric Hematology-Oncology CHU Nancy, Nancy France

^11^Service d’Hémato-Immunologie pédiatrique, Hôpital Robert Debré, AP-HP – Université Paris-Cité, Paris, France

**Supplemental Tables 1-2**

**Table S1:** **List of reagents and resources**

| **REAGENT or RESOURCE** | | | **SOURCE** | | **IDENTIFIER** | |
| --- | --- | --- | --- | --- | --- | --- |
| **Antibodies** | | |  | |  | |
| RIT1 | | | Sigma | | HPA053249 | |
| HRAS | | | Abcam | | ab86696 | |
| MRAS | | | Abcam | | ab176570 | |
| NRAS | | | Abcam | | ab77392 | |
| KRAS | | | Abcam | | ab275876 | |
| NF1 | | | Abcam | | EPR22989-68 | |
| LZTR1 | | | Abcam | | ab289965 | |
| ERK1/2 | | | Cell signalling | | #4695 | |
| P-ERK1/2 | | | Cell signalling | | #4370S | |
| MEK1/2 | | | Cell signalling | | #9126S | |
| P-MEK1/2 | | | Cell signalling | | #9154 | |
| AXL (C89E7) | | | Cell signalling | | #8661 | |
| EGFR | | | Thermo Fisher | | H9B4 | |
| β-actin | | | Novusbio | | NB600-501 | |
| Zombie violet™ Fixable Viability Kit | | | BioLegend | | #423114 | |
| **Commercial Assays** | | |  | |  | |
| RNeasy Mini kit | | | Qiagen | | #74104 | |
| QIAamp DNA Blood Mini kit | | | Qiagen | | #51104 | |
| M-PER® lysis buffer | | | ThermoFisher Scientific | | #78503 | |
| Protease Cocktail Inhibitor | | | ThermoFisher Scientific | | P8340 | |
| PhosSTOP™ | | | Roche | | #4906845001 | |
| PMSF | | | ThermoFisher Scientific | | #36978 | |
| Protein 230 kit | | | Agilent | | 5067-1517 | |
| 12-230 kDa Separation Module | | | Biotechne | | SM-W004 | |
| 66-440 kDa Separation Module | | | Biotechne | | SM-W005 | |
| Anti-Mouse Detection module | | | Biotechne | | DM-002 | |
| Anti-Rabbit Detection Module | | | Biotechne | | DM-001 | |
| Anti-Goat Detection Module | | | Biotechne | | DM-006 | |
| RePlex™ Module | | | Biotechne | | RP-001 | |
| Total Protein Detection Module for Chemiluminescence | | | Biotechne | | DM-TP01 | |
| Ras Pull-Down and Detection Kit | | | ThermoFisher Scientific | | 16117 | |
| Amaxa Human cell nucleofector kit T | | | Lonza | |  | |
| Amaxa Human SE Cell Line 4D-Nucleofector kit | | | Lonza | |  | |
| Amaxa Human SF Cell Line 4D-Nucleofector kit | | | Lonza | |  | |
| Cell Activation Cocktail (without Brefeldin A) (phorbol-12-myristate 13-acetate (25 ug/mL) and ionomycin (500 ug/mL)) | | | OZYME | | BLE9402 | |
| NucleoBond® Xtra Midi EF | | | Macherey-Nagel | | #740420.50 | |
| E.coli One Shot ® TOP10 | | |  | |  | |
| imMedia™ Kan Liquid | | | Invitrogen™ | |  | |
| CyQUANT Celle proliferation assay | | | ThermoFisher Scientific | | C35012 | |
| **Cell lines** | | | | | | |
| K562 (Human, chronic myeloid leukemia) | | | ATCC | | CCL-243™- RRID: CVCL_0004 | |
| TF-1 (Human, erythroleukemia) | | | ATCC | | CRL-2003™- RRID:CVCL_0559 | |
| RCH-ACV (Human, B-ALL) | | | DSMZ | | ACC 548- RRID:CVCL_1851 | |
| NALM-6 (Human, B-ALL) | Cytion | | | | #300297- RRID:CVCL_0092 | |
| Loucy (Human, T-ALL) | DSMZ | | | | ACC 394- RRID:CVCL_1380 | |
| TOM-1 (Human, B-ALL) | DSMZ | | | | ACC 578- RRID:CVCL_1895 | |
| Jurkat (Human, T-ALL) | DSMZ | | | | ACC 282- RRID:CVCL_0065 | |
| **Automates** | | | | | | |
| Nucleofector 2b or 4D | Lonza | | | | | |
| WOLF® Cell Sorter | NanoCellect® Biomedical Inc. | | | | | |
| Aria II FACS | BD Biosciences | | | | | |
| Bioanalyseur Agilent 2100 | Agilent | | | | | |
| Microscope Operetta | PerkinElmer | | | | | |
| **Plasmids and primers** | | | | | | |
| CMV-CG-Cas9-2A-tGFP (all-in-one Cas9-reporter) | Sigma-Aldrich | | | | custom-designed and cloned by Sigma-Aldrich | |
| *gRNA template sequences:* | | | | | | |
| gRNA LZTR1-Exon 3: | CUUUCACAUCGAACCGCAGG | | | | | (PAM sequence : AGG) |
| gRNA LZTR1-Exon 5: | UUACUCAGGGGGUUACACUG | | | | | (PAM sequence : GGG) |
| gRNA PPP1R12C (AASV1 locus) | GGGGCCACUAGGGACAGGAU | | | | | (PAM sequence : GGT) |
| *Primer sequences:* | | | | | | |
| LZTR1-Exon 3 | 5’-GCCCATATCAGTTGGGGAG | | | | | |
|  | 5’-TCGAGGTAGAGGTGGACAGC | | | | | |
| LZTR1-Exon 5 | 5’-ctctgtgtccccagagcc | | | | | |
|  | 5’-aggagggagtctAAGGGCTG | | | | | |
| PPP1R12C_Locus AASV1 | 5’-gggtcacctctcactcctttc | | | | | |
|  | 5’-aggatcctctctggctccat | | | | | |
| **URLs** | | | | | | |
| <https://depmap.org/portal/> | | | | | | |
| <https://proteinpaint.stjude.org> | | | | | | |
| <https://gnomad.broadinstitute.org/gene/ENSG00000099949?dataset=gnomad_r2_1_non_cancer> | | | | | | |
| **Deposited data** | | | | | | |
| RNAseq healthy hematopoietic samples | | | GEO repository (Accession number: GSE267628) | <https://www.ncbi.nlm.nih.gov/geo/query/acc.cgi?acc=GSE267628> | | |
| **Software and Algorithms** | | | | | | |
| R version 4.3.1 | |  | | https://www.r-project.org/ | | |
| R studio v1.6.0 | |  | | https://www.rstudio.com/ | | |
| Genome Browser | | IGV | | https://software.broadinstitute.org/software/igv/ | | |
| GraphPad Prism v10 | | GraphPad Software | | https://www.graphpad.com/ | | |
| Affinity Designer v1.9 | | Affinity | | https://affinity.serif.com/ | | |
| Synthego Crispr design Tool | |  | | https://www.synthego.com/products/bioinformatics/crispr-design-tool | | |
| ICE Analysis | |  | | https://ice.synthego.com/#/ | | |
| Broad Institute GPP sgRNA Designer | |  | | https://portals.broadinstitute.org/gpp/public/analysis-tools/sgrna-design | | |
| TrueDesign Genome Editor | |  | | https://www.thermofisher.com/fr/fr/home/life-science/genome-editing.html | | |
| ProteinPaint | |  | | https://proteinpaint.stjude.org/ | | |
| Galaxy | |  | | https://galaxy-bioinfo.aphp.fr/ | | |
| Galileo | | Integragen | | http://galileo.integragen.com/app/GeCo_App | | |
| Biology Image Analysis Software | | Single Cell Technologies | | |  | |

**ATCC:** American Type Culture Collection (Manassas, VA, USA)

**DSMZ:** Leibniz Institute DSMZ – German Collection of Microorganisms and Cell Cultures

**Table S2: Complete list of drugs tested in the *in vitro* functional screening**

| **Main category** | **Targets** | **Subcategories (compounds)** |
| --- | --- | --- |
| Pro-apoptotic agents | IAP, BCL2, MCL1, MDM2 | Birinapant, Navitoclax, Venetoclax, Obatoclax, S-63845, A-1210477, A1331852, Nutlin-3 |
| DNA-damaging agents / classical chemotherapies | Anthracyclines, antimetabolites, alkylating agents, topoisomerase inhibitors | Daunorubicin, Doxorubicin, Idarubicin, Mitoxantrone, Etoposide, Amsacrine, Topotecan, Calicheamicin, Cytarabine, Gemcitabine, Clofarabine, Fludarabine, Nelarabine, Mercaptopurine, Methotrexate, Busulfan, Carmustine, Melphalan, Mitomycin, Trofosfamide, Thiotepa, Olaparib |
| Tyrosine kinase inhibitors (TKI) | BCR-ABL, FLT3, PDGFR, FGFR, VEGFR, BTK, SRC, IGF1R | Imatinib, Dasatinib, Ponatinib, Bosutinib, Nilotinib, Midostaurin, Quizartinib, Dovitinib, Cabozantinib, Crenolanib, Gilteritinib, Erdafitinib, Linifanib, Ibrutinib, Entospletinib, BMS-509744, Sunitinib |
| RAS/MAPK pathway inhibitors | MEK, RAF | Selumetinib, Trametinib, Sorafenib |
| PI3K/AKT/mTOR inhibitors | PI3K, AKT, mTOR | Temsirolimus, Buparlisib, Ipatasertib, AZD8055, Torkinib |
| Epigenetic modulators | HDAC, DNMT, BET, Menin, p300/CBP | Vorinostat, Panobinostat, Entinostat, Nexturastat A, Givinostat, Mocetinostat, Decitabine, Thioguanine, iBET, Molibresib, A-485, VTP-50469 |
| Proteasome inhibitors | Proteasome | Bortezomib, Carfilzomib, Ixazomib, Delanzomib |
| Cell cycle / mitotic inhibitors | CDK, Aurora kinase, Plk1 | Alvocidib, Dinaciclib, Palbociclib, Ribociclib, THZ1, Barasertib, NVP-2, BI-2536 |
| Cytoskeletal signaling / HSP90 chaperone inhibitors | Microtubules, HSP90 | Vincristine, Docetaxel, HSP-990, Luminespib |
| JAK/STAT pathway inhibitors | JAK1/2, STAT | Ruxolitinib, Fedratinib, Momelotinib, BSK805, Homoharringtonine, Fludarabine |
| Metabolism / bioenergetic inhibitors | OxPhos, FOXO, DHFR | Gboxin, IACS-010759, AS-1842856, Methotrexate |
| Glucocorticoids / immunomodulators | Glucocorticoid receptor, Calcineurin | Dexamethasone, Prednisolone, Cyclosporine |
| Others / miscellaneous | Nuclear export, anti-infective | Selinexor, Eltanexor, Moxidectin |

**Supplementary Tables (provided as separate Excel files):**

**Table S3: 51 germline & somatic LZTR1 variants at diagnosis and classification**

**Table S4: Patients and controls mutations & CNV at diagnosis**

**Table S5: Hypodiploid caryotype analysis**

**Table S6: Cell Lines - RAS MAPK mutations**

**Table S7: Methods - Patients and controls - complementary analysis**
